# Identification of co-segregating *RUBCN* and *KANK1* mutations in a UK amyotrophic lateral sclerosis kindred

**DOI:** 10.1101/2025.07.23.25331426

**Authors:** Midhat Salman, Intan Bakhtiar, Alex Morris, Alexandra Rother, Simon Topp, Christopher Shaw, Bradley Smith, Jacqueline de Belleroche

**Author notes:** Correspondence to: Midhat Salman Full address: Department of Brain Sciences, Imperial College London, W12 0NN, UK. These authors contributed equally to this work (Midhat Salman, Intan Bakhtiar). Midhat Salman: Department of Forensic Medicine, University of Health Sciences Lahore, Pakistan. Intan Bakhtiar: Faculty of Health, University of Cyberjaya, Malaysia. Alexandra Rother: Max Planck Institute for Biological Intelligence, Martinsried, Germany.

## Abstract

Bioinformatic analysis of next-generation sequencing data identified novel missense variants in *RUBCN* (p.Cys517Phe) and *KANK1* (p.Pro1332Ala) that co-segregated with amyotrophic lateral sclerosis (ALS) in a United Kingdom kindred and were ultra-rare in European control populations (allele frequency <0.001%). Likelihood-based segregation analysis demonstrated significant co-segregation with disease, yielding full-likelihood Bayes factor (FLB) scores of 4.6 and 5.5 for *RUBCN* and *KANK1*, respectively, exceeding the threshold supporting pathogenicity (FLB ≥4.33). Sex-stratified penetrance modelling identified maximal FLB scores of 7.9 for both genes under a model with male and female penetrances of 0.99 and 0.01, respectively. Gene-based burden analysis of deleterious ultra-rare variants in independent ALS case-control cohorts demonstrated exome-wide significant associations with ALS (SKAT-O, p < 1 × 10^-9^). Functional studies in lymphoblastoid cell lines derived from variant carriers revealed significantly increased apoptosis and dysregulated expression of genes involved in endocytosis, LC3-associated phagocytosis, and endoplasmic reticulum stress (p < 0.05). Analysis of spinal cord tissue from ALS cases and controls showed significant upregulation of *RUBCN* (2.85-fold) and downregulation of *KANK1* (0.40-fold) (p < 0.05). Collectively, these findings implicate *RUBCN* and *KANK1* as candidate ALS genes and suggest that disruption of vesicular trafficking, and autophagy-related pathways may contribute to disease pathogenesis of ALS.

## Introduction

Amyotrophic lateral sclerosis (ALS) is a neurodegenerative disease that is characterised by the rapid progression of muscle spasticity, stiffness, and atrophy (Brown & Al-Chalabi, 2017). It typically manifests from 55 years of age and proves fatal in most patients, who succumb to respiratory failure within 3-5 years of symptom onset (Cox et al., 2010). Of the total ALS cases, approximately 10% are familial, and the rest are sporadic (Brown & Al-Chalabi, 2017). Forecasts on ALS suggest a 69% surge in cases globally from 2015 to 2040 (Arthur et al., 2016).

After the discovery of the first ALS gene, *SOD1,* the research community uncovered many other genetic contributors, such as *TARDBP*, *FUS*, and *C9orf72,* leading to a deeper understanding of molecular pathways underlying ALS pathogenesis (Arthur et al., 2016; Woollacott & Mead, 2014). To date, more than 40 genes have been identified, collectively accounting for over 80% of familial ALS (FALS) and 15% of sporadic ALS cases (Nguyen et al., 2018; Wang et al., 2023). As large multiplex pedigrees have become increasingly scarce, gene discovery efforts have shifted towards whole-exome and whole-genome sequencing of large familial and sporadic ALS cohorts. Gene-based rare-variant association methods, such as burden tests and sequence kernel association tests (SKAT), have been instrumental in identifying novel ALS genes, including *TUBA4A*, *TBK1*, *NEK1*, and *DNAJC7* (Cirulli et al., 2015; Farhan et al., 2019; Kenna et al., 2016; Smith et al., 2014). More recently, exome analysis of nearly 18,000 ALS cases and 200,000 controls identified new rare-variant risk genes, supported previously implicated genes including *DNAJC7*, and found that 12 % of cases were oligogenic (Hop et al., 2026). Nevertheless, a substantial proportion of familial ALS remains genetically unexplained and may be attributable to additional risk genes with inheritance patterns, including variable penetrance and oligogenic inheritance.

The study of large multigenerational ALS pedigrees has historically been instrumental in gene discovery; however, such kindreds have become increasingly rare, and the family sizes available have declined. Consequently, contemporary ALS pedigrees would provide valuable opportunities to investigate novel genetic risk factors and potential interactions between disease-associated genes.

In this study, we investigated kindred SM133, a multigenerational family with dominantly inherited ALS, in which two rare coding variants were identified in the corresponding genes for Rubicon, a key regulator of LC3-associated phagocytosis (LAP) and autophagic trafficking, and Kank1, a cytoskeletal regulator. Both genes have biological functions relevant to pathways implicated in ALS pathogenesis, including protein homeostasis, regulation of autophagy, intracellular trafficking, cytoskeletal organization, and neuronal maintenance (Heckmann et al., 2019; Martinez et al., 2015; Matsunaga et al., 2009; Roy et al., 2009; Sun et al., 2010; Wong et al., 2018; Zhong et al., 2009). Notably, *KANK1* has been identified as a candidate ALS susceptibility gene through integrative genomic analyses. Consistent with this, use of CRISPR-Cas9 to reproduce patient-associated KANK1 mutations in human induced pluripotent stem cell-derived motor neurons induces distal axonopathy and TDP-43 mislocalisation, a pathological hallmark of ALS (Zhang et al., 2022).

We assessed the genetic evidence through full-likelihood Bayesian segregation analysis and gene-based association testing using SKAT-O, incorporating CADD-based deleteriousness predictions for all variants. The cellular consequences of variant carriage were then investigated in patient-derived lymphoblastoid cell lines of family members. These studies were undertaken to determine whether *RUBCN* and *KANK1* represent novel ALS susceptibility genes and to explore the possibility that the disease in kindred SM133 may arise through a digenic mechanism.

## Results

### Segregation analysis of SM133 reveals mutations in *KANK1* and *RUBCN*

We performed whole exome sequencing (WES) of 17 familial ALS patients from the Imperial College London cohort, achieving a mean of 90.5% (range: 88.9–92.6%) of RefSeq coding bases covered at ≥10× depth. Whole-genome sequencing (WGS), performed in parallel on 11 of these cases, also showed very high coverage and concordance with the WES-derived variants. Seven of these cases had mutations in previously reported ALS-associated genes, including five cases (from three kindreds) with the pathogenic *C9orf72* intronic hexanucleotide expansion, as detected by repeat-primed polymerase chain reaction (PCR). One index case carried a c.1394-2del (rs1555509569) variant in *FUS* that was previously reported in 2014 (Cady et al., 2015). This mutation is predicted to cause out-of-frame skipping of exon 14 that disrupts the FUS C-terminus, similar to another previously reported ALS-causing substitution at the same site (DeJesus-Hernandez et al., 2011). A second index case harboured a c.290A>G variant, p.Lys97Arg in ENST00000397066 of the Cyclin F gene (rs1465313712 in *CCNF*). This variant was published as part of a wider study (Williams et al., 2016). The variant was found in 5/63,904 alleles in the gnomAD v3.1.2 non-Finnish European individuals who were not ascertained to have a neurological condition (‘non-neuro NFE’). Although the remaining eight index cases were free of candidate mutations in any genes previously associated with ALS, they harboured between 18 and 44 novel protein-altering variants each.

To narrow down the search for causative mutations in this subset, we focused our attention on kindred SM133, as there was a whole-exome and whole-genome sequence from the index case and an affected cousin. Additionally, the genes carrying the shared variants were expressed in the central nervous system, and their functional properties were relevant to known ALS pathogenic pathways such as autophagy and cytoskeletal formation. A novel missense single-nucleotide variant (SNV) was found in both *RUBCN* and *KANK1* genes after filtering against the gnomAD v3.1.2 ‘non-neuro NFE’ database of non-Finnish Europeans who had not been ascertained to have neurological conditions. The first *RUBCN* variant identified was a c.1550G>T transversion in exon 10 of the *RUBCN-202* (ENST00000296343, NM_014687), resulting in a missense substitution of cysteine to phenylalanine in the Rubicon protein (p.C517F). This single-nucleotide variant (SNV) was not present in the gnomAD v3.1.2 database, which included data from 67,413 individuals. The second variant, found in the same family, was a c.3994C>G transversion in exon 15 of the *KANK1-205* isoform (ENST00000382297, NM_015158, rs768866619), leading to a proline-to-alanine substitution in the Kank1 protein (p.P1332A). This SNV was likewise absent from the database, which included 67,367 individuals.

The segregation of these SNVs in the kindred SM133 (Fig. 1) was then determined by Sanger resequencing in eight individuals (Supplementary Fig. 1). The presence of both *RUBCN* c.1550G>T and *KANK1* c.3994C>G variants was confirmed in the male index case (III-2) and the second affected male cousin (III-6), as well as two female variant carriers (III-1, III-4). In addition, the *KANK1* SNV was found in III-5, a third female and sibling of III-6, who did not harbour the *RUBCN* variant. Both SNVs were heterozygotes and segregated in an autosomal dominant pattern of inheritance. Of the eight screened family members, four harboured these rare SNVs (or five for the *KANK1* mutation), and the remaining family members III-3, IV-1, and the married-in II-2a did not possess them. There were two additional obligate carriers, II-1 and II-2, with unknown status. The penetrance of these missense alleles in the SM133 kindred members appeared to be split by sex, where both male heterozygotes developed FALS compared to none of the female heterozygotes.

**Figure 1.**
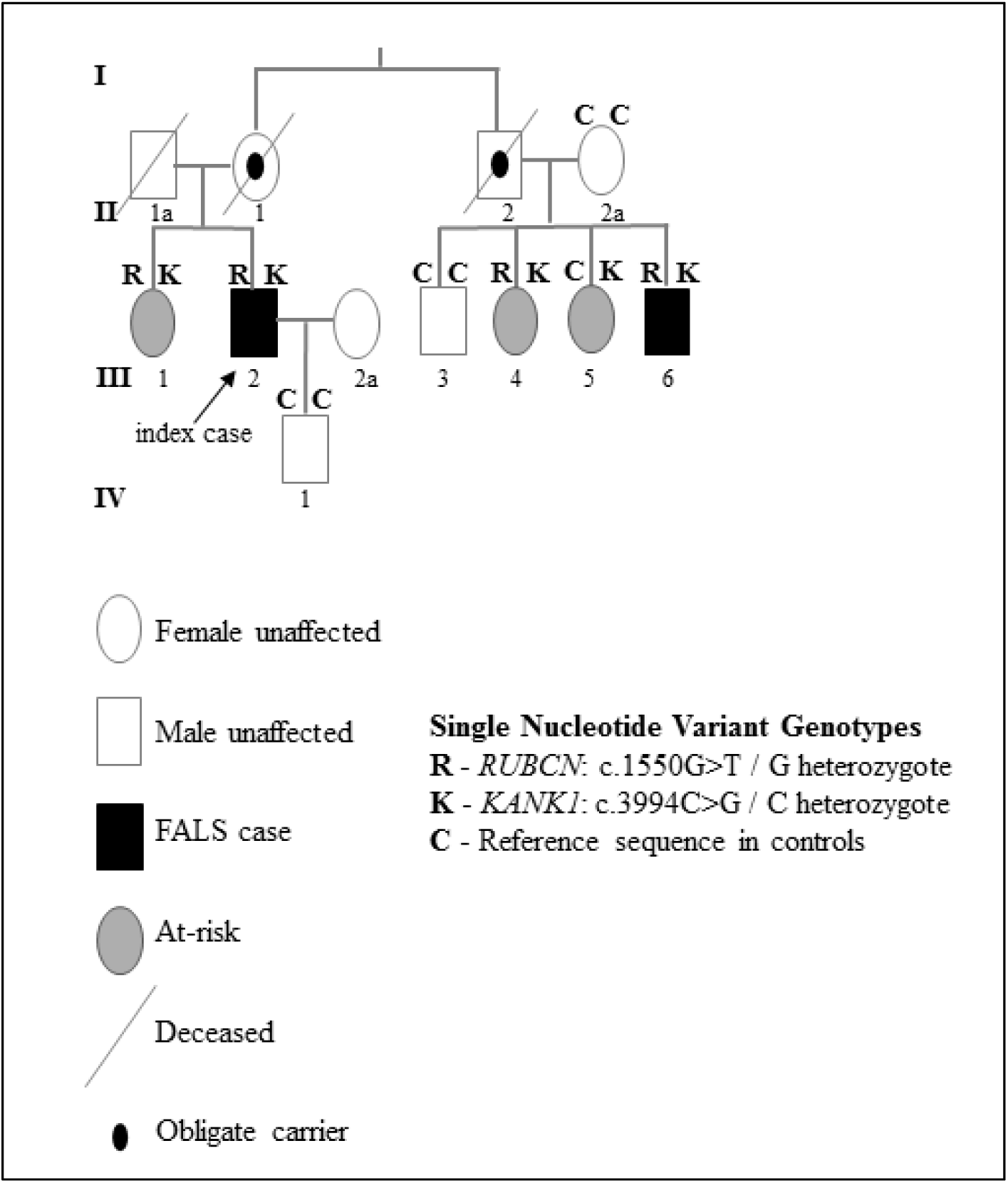
Pedigree SM133. Kindred SM133 included two affected males with familial ALS (FALS), III-2 and III-6. Blood samples and lymphoblastoid cell lines (LCLs) were available from eight family members. Four individuals were heterozygous for two rare SNVs, c.1550G>T in *RUBCN-202* (p.C517F, ‘R’) and c.3994C>G in *KANK1-205* (p.P1332A, ‘K’). In addition to the affected males, variant carriers III 1, III 4 and III 5 were all females who had not presented with ALS symptoms (III-5 had only the *KANK1* variant). The remaining unaffected individuals—III-3, IV-1, and married-in II-2a—were homozygous for the reference alleles (‘C’). The disease status of obligate variant carriers II-1 and II-2 was unknown. *RUBCN-202* (ENST00000296343) and *KANK1-205* (ENST00000382297) correspond to high-quality MANE transcripts (Matched Annotation from NCBI and EMBL-EBI).

Both *RUBCN* and *KANK1* variants were found to be either absent or have an ultra-rare MAF < 0.001 % in the European control samples described above, which is in marked contrast to the higher allele frequencies observed in the biologically related members of the SM133 kindred Cosegregation of a candidate variant with disease across a pedigree provides evidence for pathogenicity: a causal variant should be inherited by affected relatives more often than expected by chance, and largely absent in unaffected ones. In this kindred, to quantify evidence for the cosegregation of *KANK1* and *RUBCN* variants with ALS, we used the full-likelihood Bayes factor (FLB) method implemented in the R package segregatr (Thompson et al., 2003; Vigeland, 2021). This compares the likelihood of the observed pattern of variant transmission under a model where the variant is pathogenic against a model where it is neutral, producing a Bayes factor that contributes to the supporting evidence within the ACMG/AMP framework. The resulting FLB factors of *KANK1* and *RUBCN* were 5.5 and 4.6, respectively. This provides supporting evidence for pathogenicity under the Jarvik & Browning 2016 thresholds (threshold ≥ 4.33) for single families (Jarvik & Browning, 2016), based on the “Standards and guidelines for the interpretation of sequence variants” adopted by the ACMG/AMP (Richards et al., 2015).

Both male carriers of the *KANK1* and *RUBCN* variants in this kindred developed ALS, whereas none of the female carriers did, suggesting possible sex-dependent penetrance. This observation is consistent with the broader sexual dimorphism reported in ALS (Jacob et al., 2024). We therefore examined the effect of sex-specific penetrance on the FLB factor, using penetrance values ranging from 0.01–0.99 (Fig. 2). This yielded FLB values reaching a maximum of 7.9 at a male penetrance of 0.99 and a female penetrance of 0.01, corresponding to a ‘supporting’ level of pathogenicity evidence (ACMG/AMP framework) for the male cases alone.

**Figure 2.**
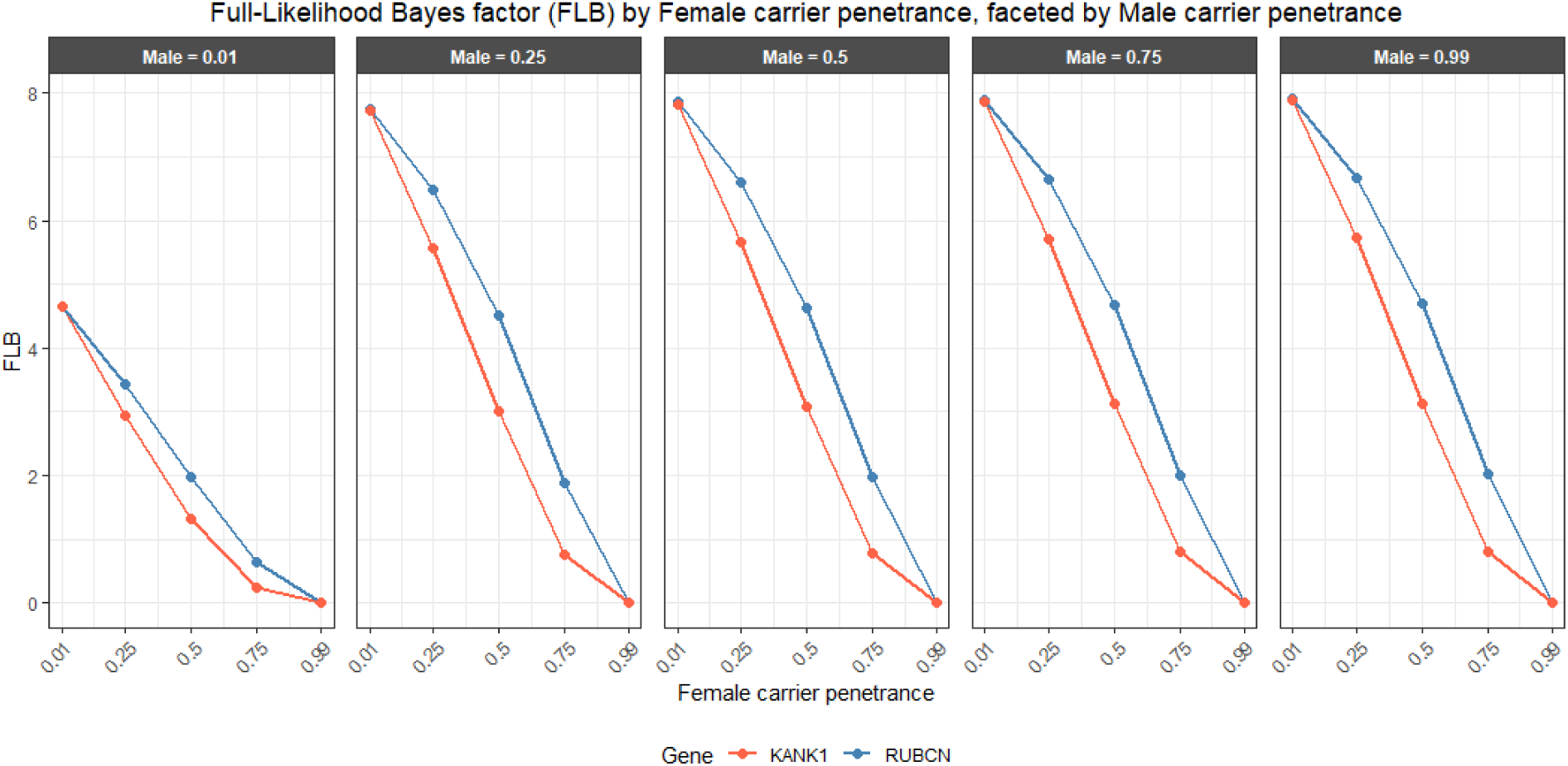
Comparison of cosegregation evidence (FLB) for RUBCN and KANK1 under varying carrier penetrance values. Panels show male carrier penetrances from 0.01 to 0.99, and each x-axis shows female carrier penetrances. Higher FLB values indicate stronger evidence in support of pathogenicity.

The pathogenicity of p.C517F in *RUBCN* and p.P1332A in *KANK1* was predicted to be probably deleterious by both PolyPhen 2 and MutationTaster. Although CADD provided PHRED scores of 17.3 and 24.5, respectively, where variants with a score ≥20 are the 1% most deleterious in the genome, neither score met a threshold of CADD ≥25 supporting deleteriousness and provided no computational evidence for this ACMG classification (Rentzsch et al., 2019) (Supplementary Table 1). We also performed multiple sequence alignments of amino acids flanking the missense variants to reveal their conservation across species, indicating their functional significance. The reference amino acids, p.Cys517 in Rubicon and p.Pro1332 in Kank1, along with their flanking residues, were found to be conserved in their evolutionary orthologs: Cys 517 was conserved across eutherian mammals, whereas Pro 1332 was further conserved between humans and insects (Supplementary Fig. 2).

### Gene-level burden testing shows a significant association of *KANK1* and *RUBCN* with ALS

Since rare variants are present in only a small number of people, they may not show a significant association with disease. For a given gene, burden tests detect rare variant effects by evaluating the cumulative association of multiple rare variants with disease in cases and controls. For *KANK1* and *RUBCN*, we performed burden tests using the aggregate of pathogenic rare missense SNVs with a CADD score ≥ 20 in each gene (the Cys517Phe variant in *RUBCN* was excluded due to a CADD score < 20). The cases included our FALS cohort and the ALSdb3 and MinE datasets, totalling 7,696 ALS patients with variants absent or present at <0.001% frequency in controls. The control dataset 1included singleton missense variants with allele frequencies <0.001%, enabling detection of evolutionarily recent alleles, critical for traits under purifying selection (Povysil et al., 2019). Consequently, collapsing analyses often produce the strongest signals from the rarest variants.

To implement these tests, a combined control sample of 53,424 individuals was used, consisting of 1,832 MinE ALS controls and 51,592 non-Finnish European ‘non-neurological’ controls from gnomAD who had not been ascertained with a neurological condition (see Methods). Assessing concordance between these control datasets is important, as differences in population composition can systematically alter allele frequencies and confound association analyses. To assess potential population stratification between the gnomAD and MinE control datasets, we compared single-nucleotide variants (SNVs) with minor allele frequencies (MAFs) >0.05% in RUBCN and KANK1. Allele frequencies were highly concordant between datasets (R² = 0.999), indicating minimal population stratification of the control populations.

Out of the 7,696 ALS cases, seventeen and twenty-five missense variants were found in *RUBCN* and *KANK1,* respectively. Moreover, three doubletons were found in the patient group: two in *RUBCN* (A504V and R813Q) and one in *KANK1* (N1156T). None of the above variants were found in the control group. In *KANK1,* the G1002R and P1332A variants were found in one case and a control. The control samples had five *RUBCN* and twenty *KANK1* ultra-rare variants (Fig. 3). The association of the SNVs in each gene with ALS was estimated by the SKAT-O binary robust tests implemented in R, which estimated a p-value < 1×10^-9^ for both genes (supplementary Fig. 3). This is significant relative to a threshold for whole-exome sequencing (WES) of *p* = 2.5 × 10^-7^ (Fadista et al., 2016). As an orthogonal test of rare-variant burden, we applied Fisher’s exact test to carrier counts at each locus. This corroborated the SKAT-O findings, demonstrating significant enrichment of variant carriers compared with population controls for *RUBCN* (OR = 24.99, 95% CI: 8.95–86.25, *p* = 1.1 × 10^-^¹²) and *KANK1* (OR = 9.1, 95% CI: 4.84–17.58, *p* = 3.8 × 10^-^¹²).

**Figure 3.**
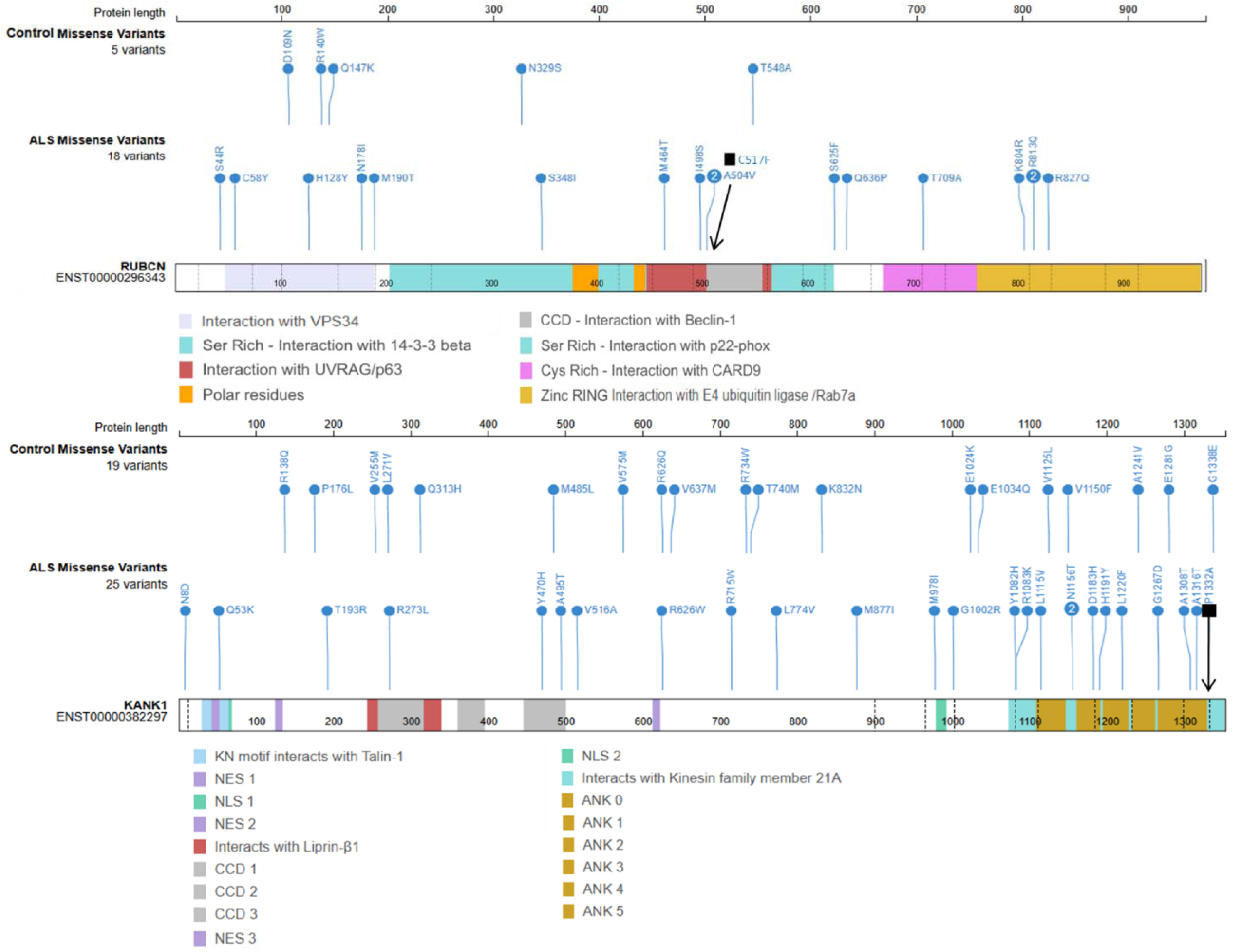
Missense SNVs in *RUBCN* and *KANK1*. Missense SNVs found with a CADD score > 20 in 7696 ALS patients from the MinE and ALSDB3 databases that were selected for an ultra-rare frequency of 0 to 0.001% in 53,424 neurological controls (MinE controls and the gnomAD non-Finnish European individuals, who were diagnosed as not having a neurological disease); the black square and arrows indicate the corresponding SNVs found in kindred SM133. **(a)** Ultra-rare ALS SNVs in *RUBCN*-202 showing C517F (black arrow) and a further 17 variants, including the two doubletons at A504V and R813Q. **(b)** 25 ultra-rare ALS SNVs in *KANK1*-206, showing P1332A (black arrow) and the doubleton at N1156T. The 5 and 19 ultra-rare control variants found in *RUBCN* and *KANK1*, respectively, were selected using the same criteria. The vertical dashed lines show the exon boundaries. Abbreviations: Ankyrin domain (ANK) repeats; Coiled-Coil Domains (CCD); Nuclear Export Signal 1 (NES): Nuclear Localization Signal 1 (NLS); RPIP8, Unc-14 And NESCA domains (RUN).

### Transcript analysis identifies dysregulation of the endocytosis, ER stress, and autophagy pathways

The *RUBCN* gene encodes the Rubicon protein, which is widely expressed in most tissues of the body and is abundant in both cerebral cortex and spinal cord neurons (Beltran et al., 2019; Wong et al., 2018). Rubicon is localised to the late endosome and lysosome compartments (Matsunaga et al., 2009). It consists of multiple domains that interact with other proteins of canonical and non-canonical autophagy and endocytosis pathways (Sun et al., 2010; Yang et al., 2012). These features enable Rubicon to play a role in multiple signalling pathways: Rubicon down-regulates the maturation steps in the canonical pathways of autophagy and endocytosis (Matsunaga et al., 2009; Sun et al., 2010; Zhong et al., 2009). Conversely, Rubicon is an essential positive regulator of the NADPH oxidase complex in the non-canonical pathway of LC3-associated phagocytosis (LAP) (Martinez et al., 2015) and in LC3-associated endocytosis (LANDO) (Heckmann et al., 2019). Kank1, KN Motif and Ankyrin Repeat Domains 1, attenuates the remodelling of cytoskeletal actin fibres during neurite outgrowth and lamellipodia formation, with a role also in cell polarity and migration (Gee et al., 2015; Roy et al., 2009). Kank1 cross-talks between focal adhesions and cortical microtubules by physically linking the two macromolecular assemblies with the microtubule-stabilising complex. It does this via several proteins, including KIF21A (Bouchet et al., 2016; Rafiq et al., 2019).

Therefore, we performed functional studies using cellular models to investigate the influence of *RUBCN* and *KANK1* mutations on the relevant cellular pathways. We investigated the effects under basal conditions (without drug treatment) and cell stress by gene expression profiling of pathway markers using qPCR. We chose lymphoblastoid cell lines (LCLs) as a cellular model since they have previously been used to identify altered signalling pathways in neurological disorders (Annesley & Fisher, 2021). We used eight LCLs derived from the SM133 kindred (Fig. 1) to compare LCLs containing both *RUBCN* and *KANK1* mutations with wild-type LCLs. *RUBCN* and *KANK1* mutations in LCLs were confirmed using Sanger sequencing. The following pathway markers were selected for investigation: (1) *RAB7A* for endocytosis, (2) *NOX2* for LC3-associated phagocytosis (LAP). We measured three autophagy pathway transcripts: (3a) *RUBCN*, as well as two components of the PI3KC3 complex, (3b) *VPS34* and (3c) *UVRAG*. Three markers of the ER stress pathway, (4a) *DNAJC10*, (4b) *HSPA5*, (4c) *XBP1,* and finally, (5) *KANK1* for cytoskeletal organization. Cell stress was induced in LCLs to create a model that mimics the cellular conditions of ALS by using three drugs (rapamycin, tunicamycin, and thapsigargin) for activating autophagy and ER stress.

Under basal conditions, the *KANK1* and *RUBCN* mutations caused a significant dysregulation in the expression of the following marker genes in LCLs (Fig. 4): for endocytosis, downregulation of *RAB7A* (fold-change (*FC*) = 0.38, p-value = 0.02) and for LC3-associated-phagocytosis (LAP), downregulation of *NOX2 (FC* = 0.43, *p* = 0.01). Of the ER stress transcripts, *DNAJC10* was downregulated (*FC* = 0.53, *p* = 0.03) and *XBP1* was upregulated (*FC* = 2.83, *p* = 0.006) (Supplementary Table 2). Overall, these significant changes suggest that the mutations dysregulated endocytosis, LAP, and ER stress.

**Figure 4.**
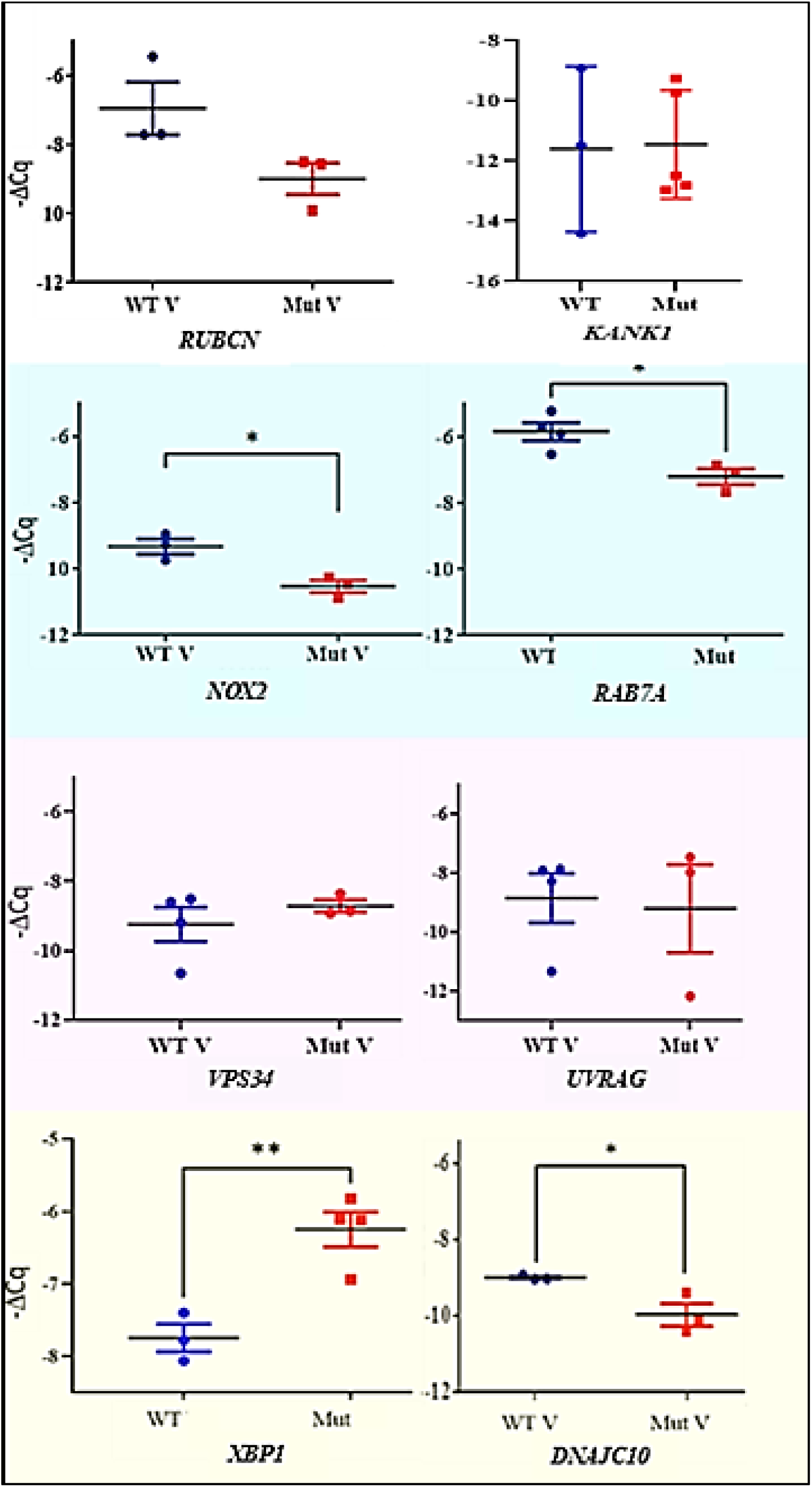
Effect of the C517F and P1332A mutations on pathway transcript levels. Effect of the *RUBCN* C517F and *KANK1* P1332A mutations on basal transcript levels in lymphoblastoid cell lines (LCLs) from variant carriers and wild-type individuals in the SM133 kindred. Data are presented as –ΔCq (–ΔCt) values with means ± SEM. The top row shows transcript levels of *RUBCN* and *KANK1*. The blue panel includes transcripts of LAP and endocytosis marker genes, the pink panel highlights autophagy transcripts, and the yellow panel shows ER stress respons genes. Significant mutation effects on *RAB7A*, and *XBP1* expression were observed in well containing medium alone, whereas significant effects on *NOX2* and *DNAJC10* expression were detected in wells supplemented with 0.4% DMSO (vehicle), which was present for 4 hours. Expression levels were normalised to *ACTB*. Bars represent mean ± SEM: blue for wild-type, red for mutant. Statistical analysis was performed using unpaired Student’s t-tests (\**p* < 0.05, \*\**p* < 0.01). WT – wild-type; Mut – mutant; V – vehicle control for basal expression.

Rapamycin (R) induces autophagy by inhibiting the mammalian target of rapamycin (mTOR). In both wild-type and mutant LCLs, R treatment caused a significant downregulation of *RUBCN* mRNA expression compared to that in the untreated LCLs (Supplementary Table 3). In the wild-type LCLs, R also caused a significant downregulation of mRNA expression for the autophagy marker gene *VPS34*, the endosomal pathway marker *RAB7A,* as well as the ER stress marker *DNAJC10,* compared to untreated LCLs. In contrast, for the corresponding mutant LCLs, the effect of R on these genes was not evident, suggesting that the mutation may have disabled the downregulating effect of rapamycin.

### The *RUBCN* and *KANK1* mutations significantly increase apoptosis

The FITC-Annexin V assay in conjunction with cell sorting was used to examine the effect of *RUBCN* p.C517F and *KANK1* p.P1332A mutations on lymphoblastoid cell lines. The effects on cell survival were measured under basal and cell stress conditions. ER stress was induced by treatment with tunicamycin (TN) or thapsigargin (TG).

Compared to wild-type LCLs, the mutations showed a significant increase in total apoptosis when cultured in vehicle only (*p* < 0.05) (Fig. 5). Drug treatment itself gave rise to significantly elevated total apoptosis levels in cell lines (repeated measures ANOVA, *p* < 0.05), indicating the induction of the relevant pathways.

**Figure 5.**
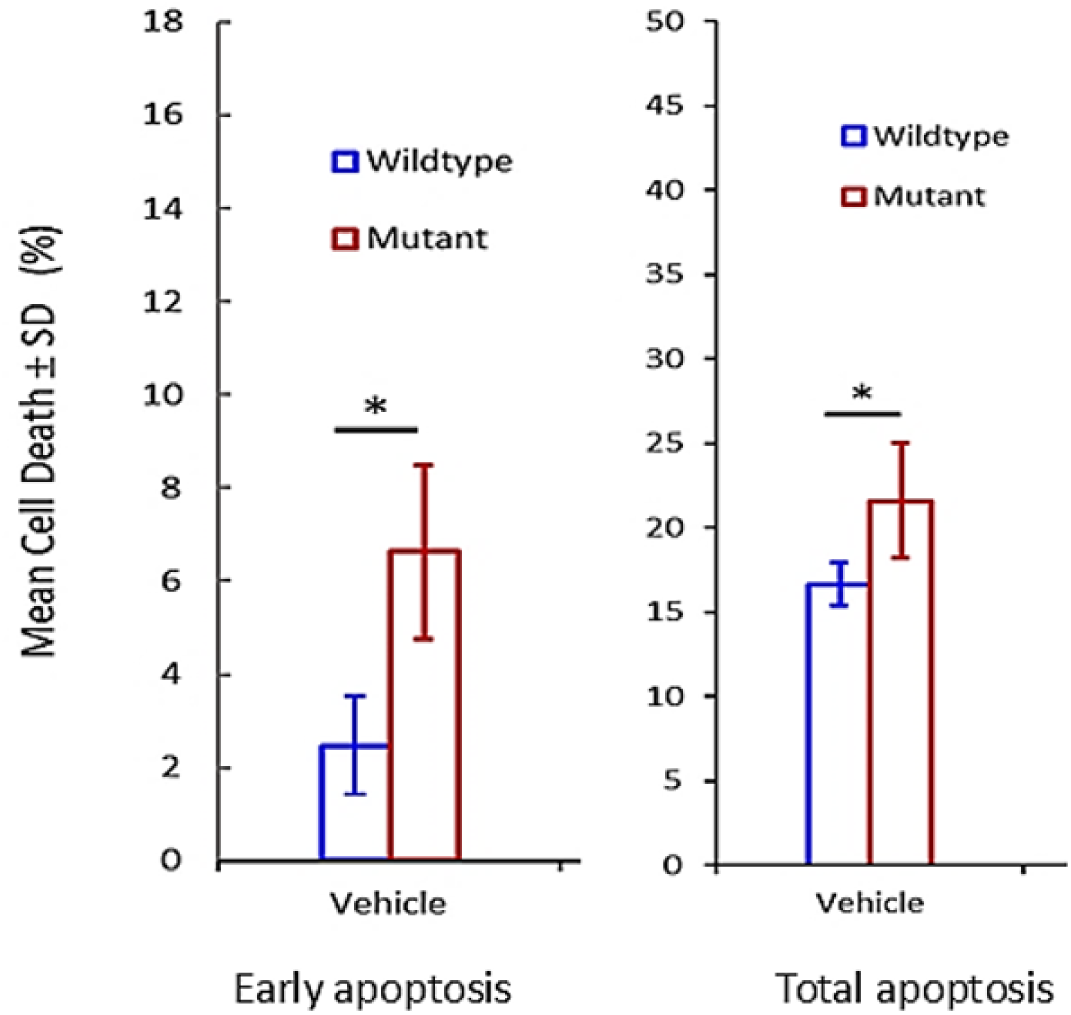
Annexin V assay for cell survival. Means and standard deviations (SD) of % cell death in early apoptosis and total apoptosis detected by flow cytometry with the annexin V assay. The measurements were of wild-type (blue, *n* = 3) or mutant lymphoblastoid cell lines (red, *n* = 5) treated with vehicle. Apoptosis levels of mutant versus wildtype cells were compared using unpaired two-tailed t-tests (*p* < 0.05 is marked by *).

### *RUBCN* and *KANK1* are involved in sporadic ALS Pathogenesis

Based on our findings in LCLs, we investigated potential alterations in *RUBCN* and *KANK1* expression levels, as well as markers of pathways involved in the action of these genes, in sporadic ALS (SALS) pathology. For this purpose, human spinal cord post-mortem samples derived from SALS cases were compared to age-matched healthy control samples available in the laboratory.

Using qPCR, we found a significant increase in *RUBCN* mRNA expression (*FC* = 2.85, *p* = 0.0003) (Fig. 6). Similarly, *KANK1* mRNA expression level was significantly decreased (*FC*=0.81, *p* =0.023) in SALS cases compared to controls. Additionally, we found a corresponding significant decrease in the mRNA expression of the binding partner of KANK1, *KIF21A*, in SALS cases compared to controls (*p*<0.0001) (Fig. 7). The KANK1/KIF21A interaction is evolutionarily conserved in different species (Weng et al., 2018) and is also important for cell migration and polarity (Gee et al., 2015), where KIF21A inhibits the formation of microtubules in the cortical regions of cells (van der Vaart et al., 2013).

**Figure 6.**
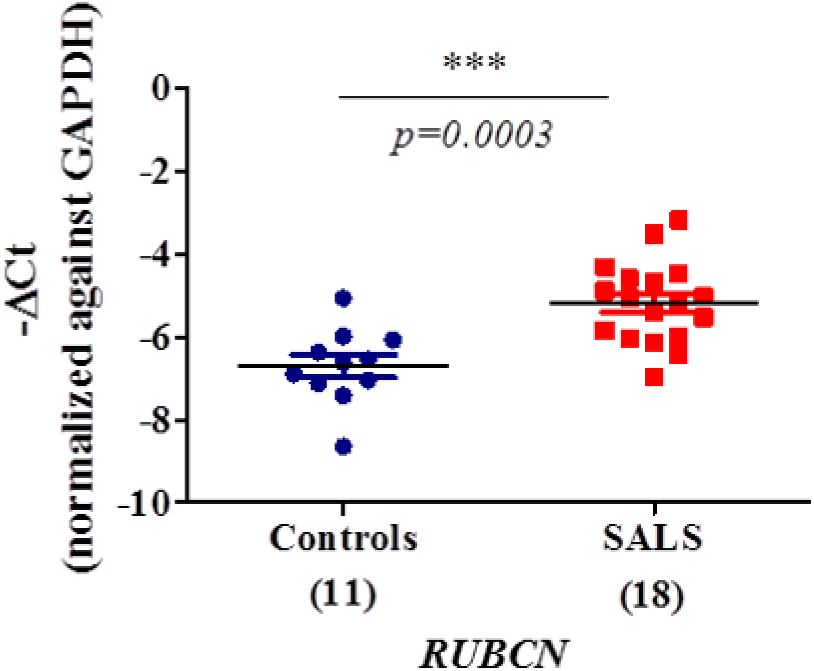
Levels of *RUBCN* mRNA in spinal cord. *RUBCN* mRNA expression levels in th spinal cord post-mortem samples from SALS cases (red dots) and controls (blue dots). The number of samples analysed is given in brackets below the X-axis. The gene expression value were normalised to *GAPDH* or *ACTB* and are given as -ΔCt where the black bars show means and coloured bars, SEMs. The data were normally distributed as tested by the D’Agostino and Pearson normality test and were compared using the unpaired Student’s t-test (\*\*\**p* < 0.0005).

**Figure 7.**
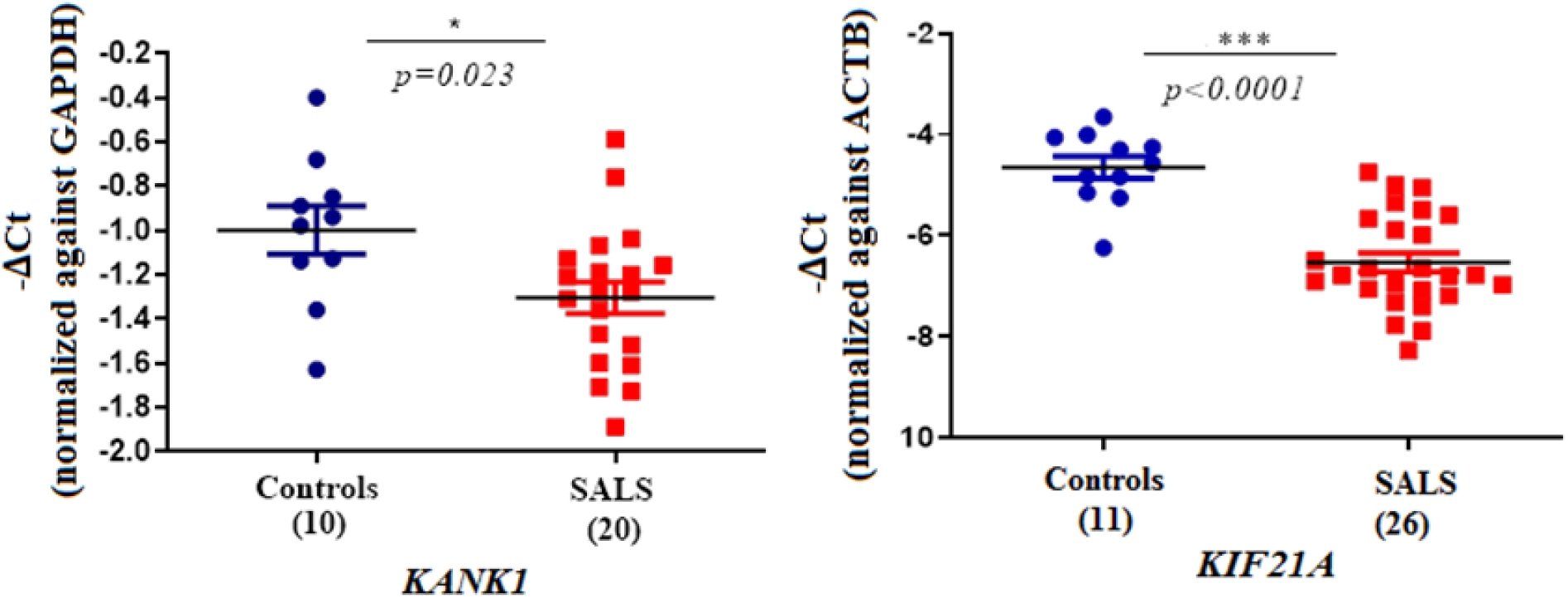
Levels of *KANK1* and *KIF21A* mRNA in spinal cord. Quantitative PCR analysis of spinal cord. Using unpaired t-test, normalised -ΔCt values showed a significant decrease both of *KANK1* total transcript levels in SALS compared to controls, and of *KIF21A* in SALS individuals compared to controls (\**p* < 0.05, \*\*\**p* < 0.0005). Error bars represent standard error of means (S.E.M).

## Discussion

We identified the novel mutations in *RUBCN* (c.1550G>T) and *KANK1* (c.3520C>G) using whole-exome data from the Imperial College FALS cohort. Both mutations are ultra-rare in European control datasets (<0.001%) but were identified in at least seven carriers and observed to cosegregate with disease in two affected male cousins in this three-generation family. The presence of mutations in these two genes is consistent with the oligogenic basis of FALS, recently estimated at approximately 12% of ALS-associated variants (Hop et al., 2026). When analyzed using a full-likelihood Bayes factor method, implemented via the segregatr R package (Bulmer et al., 2025; Hop et al., 2026; Vigeland, 2021), FLB values of 4.6 and 5.5 were obtained for the two variants, respectively. In the framework of Jarvik and Browning, an FLB value exceeding the threshold of 4.33 for a single family can be considered supporting evidence for cosegregation of the variant with FALS (Jarvik & Browning, 2016). Based on male penetrance values from 0.25 to 0.99 and a minimal female penetrance of 0.01, the FLB reached a maximum that remained relatively stable for both genes (7.8–7.9; Fig. 2), providing supporting evidence for cosegregation in the two affected male carriers. In contrast, as female penetrance increased, FLB declined markedly for both genes, consistent with the presence of two unaffected female carriers in the *RUBCN* and three in the *KANK1* pedigrees. For this kindred, its modest size together with incomplete penetrance, limited the maximum attainable FLB and consequently, the strength of the cosegregation evidence.

An optimized combined burden and sequence kernel association test (SKAT-O) of aggregated ultra-rare coding variants across the *RUBCN* and *KANK1* genes also demonstrated an association with ALS, exceeding the exome-wide significance threshold (p < 2.5 × 10^-7^). The rarity of these minor alleles (carrier counts ranging from 5 to 25) resulted in relatively wide 95% confidence intervals for the odds ratios from Fisher’s exact test, a known limitation of rare-variant analyses. Nevertheless, the lower bounds of the confidence intervals (8.95 for *RUBCN* and 4.84 for *KANK1*) remained well above 1, indicating robust statistical evidence for association despite the imprecision of the effect estimates. For missense variants with a MAF greater than 0.05% in the control datasets, the near-perfect concordance of MAF (R² = 0.999) between the MinE and gnomAD cohorts argues against substantial population stratification or systematic differences in ancestry.

The functional impact of these two missense variants on the viability of the EBV-transformed LCLs, derived from cases and controls from the kindred, was shown by a significant increase in total apoptosis in the mutant carrying LCLs versus non-carrier unaffected family members. While the FITC-annexin V assay is commonly used to detect apoptosis, incorporating complementary methods such as DAPI staining may better distinguish apoptotic from necrotic cell death and refine mechanistic interpretation.

Furthermore, our qPCR measurements of the transcripts revealed a significant upregulation of *XBP1* and a downregulation of *DNAJC10* in mutant LCLs compared to wild-type LCLs under basal conditions. *XBP1* is a component of the IRE1-mediated unfolded protein response, and *DNAJC10* functions as an endoplasmic reticulum co-chaperone in the ERAD pathway. Both markers have previously been reported to be upregulated in the lumbar spinal cord of patients with sporadic ALS, indicating the activation of the UPR pathway (Montibeller & de Belleroche, 2018). A significant downregulation of *DNAJC10* in our LCLs with DMSO added for 4 hours, therefore, points to the possible disruption of the UPR pathway. UPR has already been widely associated with neuronal dysfunction in neurodegenerative diseases, including ALS (Karademir et al., 2015; Montibeller & de Belleroche, 2018). Moreover, *DNAJC7*, a gene paralogous to *DNAJC10*, has been associated with ALS (Farhan et al., 2019; Hop et al., 2026) where p.Arg156Ter led to decreased protein expression in an ALS case, which is in the same direction as the decreased *DNAJC10* expression in our mutant LCLs. These findings suggest that the disturbed expression of components of the UPR machinery could contribute to increased cell death in the mutation-containing LCLs.

The gene product of *NOX2* interacts with the C-terminal segment of Rubicon (Supplementary Fig. 4). Under basal conditions with DMSO added for 4 hours, the mutant LCLs showed significantly lower mRNA expression levels of *NOX2*, a marker gene for the LAP pathway. Since Rubicon recruits NOX2 protein for LAP (Martinez et al., 2015) by binding it to amino acids 558-625 (Yang et al., 2012) the nearby p.C517F mutation may have led to the associated downregulation of the *NOX2* transcript levels. This, in turn, could impair the LAP pathway by decreasing the level of reactive oxygen species production (Supplementary Fig. 5). The deleterious effects of this mutation may be relevant to microglia in the human CNS. In a murine model of multiple sclerosis, deletion of *Atg7* in microglia led to impaired LAP, which failed to clear tissue debris and caused persistent neuroinflammation (Berglund et al., 2020). This stood in contrast to the controls, where functional LAP carried out efferocytosis that curbed both disease progression and neuroinflammation. Similarly, in Alzheimer’s disease, Heckmann et al. found that a deficiency of microglial Rubicon in the related pathway, LC3-associated endocytosis (LANDO), also led to accelerated neurodegeneration with elevated neuroinflammation (Heckmann et al., 2019). Moreover, in another study, very low NOX2 levels were shown to promote autoimmune neurodegeneration (Sorce et al., 2017). Together, these studies suggest that the decreased *NOX2* expression resulting from the p.C517F mutation in LAP may also lead to defective efferocytosis and impaired anti-inflammatory responses. Confirmation of this will require further studies.

Similar to NOX2, Rab7 also interacts with the C-terminal segment of Rubicon (Supplementary Fig. 4) and is a marker gene for the endosomal pathway. These missense variants were also associated with a significant decrease in mRNA expression levels of *RAB7A* under basal conditions in the present study. Rubicon is a negative regulator of endosomal maturation. Active GTP-bound Rab7, upon binding to Rubicon, releases the UVRAG-containing PI3KC3 complex from Rubicon sequestration and allows endosomal maturation and subsequent lysosomal degradation of its cargo (Lin & Zhong, 2011; Sun et al., 2010). The significant downregulation of the *RAB7A* mRNA expression indicates a possible hindrance in endosomal maturation, which could have resulted from binding to disrupted Rab7A on the RH domain of p.C517F Rubicon. A previous report of a frameshift mutation in *RUBCN,* associated the gene with autosomal recessive spinocerebellar ataxia-15. The mutation, p.Ala875ValfsX146, is adjacent to the C-terminal domain that binds to Rab7A and impairs the endosomal localisation of Rubicon (Assoum et al., 2013). Further studies are required to determine whether the p.C517F *RUBCN* disrupts the interaction of Rubicon with Rab7A and its endosomal localization.

Although DMSO concentrations exceeding 1% are known to induce apoptosis and oxidative stress, concentrations below 0.5% are generally considered physiologically non-disruptive (Smith et al., 2016; Trubiani et al., 1996). Interestingly, our results suggest that even such low-level exposure can act as a mild stressor capable of unmasking some transcriptional effects of disease-associated mutations. In our mutant cell lines, treatment with 0.4% DMSO induced notable alterations in pathways linked to *DNAJC10* and *NOX2*, which were absent under DMSO-free basal conditions. This observation implies that the mutations sensitize cells to minimal perturbations, thereby unmasking subtle regulatory vulnerabilities that could contribute to their pathogenic behaviour under stress-related conditions.

The Rubicon site containing the p.C517F mutation interacts with UVRAG and Beclin-1 (supplementary Fig. 4). Since both of these proteins are components of the class III phosphatidylinositol 3-kinase (PI3KC3) complex, which is involved in autophagy, we hypothesized that the mutation might affect the binding and/or expression of either or both proteins with Rubicon. Unexpectedly, the mutation had no effect on the expression of *BECN1,* and *UVRAG*. This could be explained by the ability of Rubicon to maintain its binding to the PI3KC3 complex via Vps34, which has multiple interactions with Beclin1 and UVRAG (Supplementary Fig. 4) (Martinez et al., 2015; Sun et al., 2010).

Rubicon shares high sequence homology with a previously reported ALS-associated protein, Pacer (also known as Rubicon-like), which acts as a positive regulator of autophagosome maturation, in contrast to the inhibitory activity of Rubicon (Beltran et al., 2019). Like Pacer, Rubicon is also expressed in spinal cord neurons (Beltran et al., 2019), signifying its importance in these cells. A recent study on astrocytes revealed a downregulation of *RUBCN* expression in ALS, which raised the possibility that *RUBCN* is a protective gene whose effect is lost in ALS (Ziff et al., 2022). Similarly additional evidence that *KANK1* is a risk factor for ALS was previously obtained using CRISPR-Cas9 introduced mutations in motor neurons derived from induced pluripotent stem cells (iPSCs) to recapitulate the functional effect of exonic nonsense mutations. These motor neurons undergo increased nuclear fragmentation and apoptosis. Furthermore, they displayed significantly shorter neurite and branch lengths and mislocalization of TDP-43 from nucleus to cytoplasm, a hallmark of most ALS cases (Zhang et al., 2022).

Our qPCR expression studies on the involvement of *RUBCN* in sporadic disease, using post-mortem lumbar spinal cord samples from patients with SALS and controls, showed a substantial upregulation of *RUBCN* mRNA levels in SALS. These results confirmed the previous findings of Beltran et al., which were obtained from a smaller number of samples (two controls and six SALS), showing a corresponding increase in Rubicon protein expression (Beltran et al., 2019). Our threefold upregulation of *RUBCN* mRNA in SALS suggests a hypothesis for a disease process involving a shift from macro-autophagy to LAP, which is consistent with Rubicon’s inhibition of macro-autophagy and activation of noncanonical autophagy pathways, such as LAP and LANDO (Heckmann et al., 2019; Martinez et al., 2015). We observed a significant downregulation of *KANK1* and *KIF21A* mRNA in SALS cases. KIF21A and KANK1 protein interactions are necessary for the development of the dendritic spine and synaptic plasticity (Sun et al., 2025). Downregulation of both proteins indicates defects in these processes.

The ACMG/AMP framework for variant pathogenicity specifies four levels of strength of evidence: supporting, moderate, strong, and very strong. The following outlines the evidence obtained in this study, appraised against these criteria. The missense variants of *RUBCN* and *KANK1* identified in the kindred SM133 were very rare in population reference datasets, supporting moderate evidence of pathogenicity based on allele frequency alone. Segregation analysis within this kindred, applying the full likelihood Bayesian approach, yielded factors for both variants that exceeded the established single-family threshold for supporting evidence. Sequence conservation flanking each variant showed that each was evolutionarily constrained, lending biological plausibility. Gene-level burden testing revealed a statistically significant excess of rare variants in affected cohorts, providing strong evidence in support of pathogenicity for both genes. Functional characterization across three complementary assays provided convergent support for pathogenicity: transcriptomic pathway analysis in digenic carrier-derived lymphoblastoid lines met the criteria for moderate functional evidence, and viability and apoptosis assays in the same cell lines reinforced this signal. The main shortfall was the absence of a neuronal or isogenic cell model with rescue experiments. For *KANK1*, the findings of Zhang et al., derived from a neuronal model system (Zhang et al., 2022), adds strong functional support for the pathogenicity of this gene. Finally, the significant dysregulation of *RUBCN* and *KANK1* gene expression in spinal cord tissue from SALS patients, confirmed their involvement in this disease, without however measuring the effect of specific variants. Overall, the functional evidence was considered to meet a moderate level for *RUBCN* and *KANK1*, pending future validation in a motor neuron model. Each piece of genetic and functional evidence, as summarized above, was graded using the hierarchical labels of the Tavtigian et al. framework (Tavtigian et al., 2020). Here, levels labeled as supporting, moderate, and strong evidence carry increasing weight. The cumulative classification falls in the ‘likely pathogenic’ range for *RUBCN*, and near the uppermost boundary of likely pathogenic for *KANK1*. A second independently cosegregating family would add support for the ‘pathogenic’ classification of these variants. Future studies may benefit from organoid models that better recapitulate tissue architecture and the cellular interactions present in ageing neural tissue. Histological analysis of subcellular protein localization may also provide further insight into ALS pathogenesis and its relationship with age-dependent disruption of proteostasis and autophagy.

## Conclusion

In summary, the p.C517F variant in *RUBCN* and the p.P1332A variant in *KANK1* were absent in two large control cohorts but were enriched in eight consanguineous kindred members, with minor allele frequencies of 28.6% and 35.7%, respectively. Both variants were significantly associated with disease status within the kindred, and rare missense variants in both genes showed an exome-wide association with ALS in Northern European samples using SKAT-O analysis. Functional assays in lymphoblastoid cell lines from variant carriers revealed dysregulation of non-canonical autophagy, unfolded protein response, and endocytosis. These disruptions were accompanied by significantly elevated apoptosis, suggesting that impaired stress responses and trafficking mechanisms may collectively contribute to the mutation-driven cell death. In conjunction with transcriptomic data from sporadic ALS spinal cord cases, our results strongly support *RUBCN* and *KANK1* as novel ALS-associated genes. The pleiotropic impact of these variants on aging-relevant cellular pathways is illustrated in Fig. 8, highlighting their potential role in driving age-associated neurodegenerative susceptibility.

**Figure 8.**
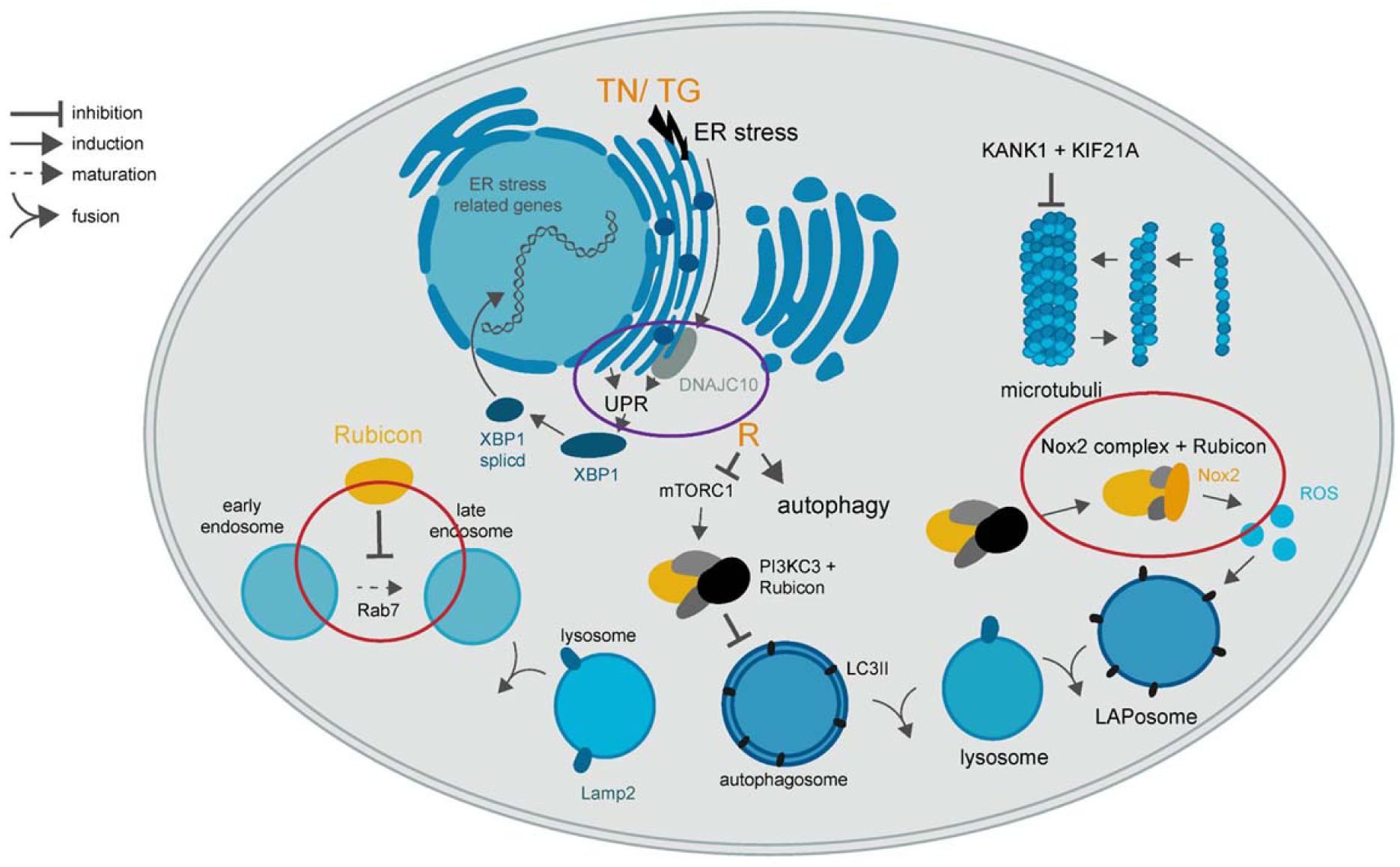
Overview of the changes occurring inside the cell from the p.C517F *RUBCN* and p.P1332A *KANK1* mutations. In this study, we assessed the effects of the p.C517F *RUBCN* and p.P1332A *KANK1* mutations on autophagy, endocytosis, LC3-associated phagocytosis, and ER stress pathways. Rapamycin (R) was used to induce autophagy and tunicamycin (TN) or thapsigargin (TG) were used to induce ER stress. The two mutations jointly affected th expression of the *Rab7*, *NOX2*, *DNAJC10*, and *XBP1*, affecting endocytosis, LC3-associated phagocytosis, and UPR pathways. We propose that these cellular changes jointly led to cell death. Circles indicate the genes whose expression was affected by the mutations.

## Materials and Methods

### Whole Exome and Genome Sequencing

DNA was isolated from blood cells of 17 patients (13 index cases) from the Imperial College London Familial ALS cohort, and exome capture was performed using standard protocols with Roche NimbleGen SeqCap EZ Exome V3 probes. The samples were exome sequenced as part of the first international FALS exome consortium, which led to the association of rare variants in *TUBA4A* with ALS (Smith et al., 2014). Paired-end sequencing was carried out at the Guy’s Hospital Genomics Facility, London, on an Illumina Hi-Seq 2000 to produce 100bp reads. In parallel, DNA from 11 of the patient lymphoblastoid cell lines (LCLs) with nine index cases was also whole-genome sequenced at Massachusetts General Hospital, Boston, producing 150-bp paired-end reads. Reads were assembled to the hg19 human reference genome using NovoCraft Novoalign. Variants were called with samtools-mpileup, normalised with bcftools norm, and annotated using Annovar and additional custom Perl scripts. Variants were quality filtered at DP ≥ 10, GQ ≥ 90, and MQ ≥ 50. Variants were excluded if they were present in any of the following population datasets: the 1000 Genomes Project, 670 local control exomes, UK10K exomes, Exome Variant Server, ExAC, Greater Middle Eastern Exomes, or the non-neurological subset of gnomAD. Variants were further excluded if a manual inspection in the Integrated Genomics Viewer suggested they were false-positive variant calls. Potential effects on splicing were assessed for variants within 25 bp of a known splice site by ADA, randomForest, Spidex, NetGene2 and GeneSplicer software. Synonymous, 5′ or 3′ UTR and intronic variants were excluded from consideration in this analysis unless they were predicted to alter splicing by two or more independent algorithms, as well as loci with poor coverage in gnomAD v2.1.1 (i.e. a read depth ≥10x in <60,000 non-neurological individuals).

### Samples

This study was approved by the Riverside Research Ethics Committee, The NRES committee Fulham, and the Genetics of Motor Neurone Disease/Amyotrophic Lateral Sclerosis. REC reference: 12/LO/0902. NRES Committee Yorkshire and the Humber. Gene expression in Motor Neurone Disease/Amyotrophic Lateral Sclerosis. Reference: 12/YH/0281, and the study was conducted according to the guidelines. Patients with ALS were diagnosed according to the El Escorial diagnostic criteria defined by the World Federation of Neurology (Brooks, 1994; Brooks et al., 2000). Consent was obtained from the participants in accordance with the Declaration of Helsinki.

SM133 FALS samples: Blood samples or buffy coats were available from eight individuals of the SM133 kindred. At the time of ascertainment, two males were diagnosed with FALS (III-2 and III-6), while the remaining six did not present with ALS symptoms (III-1, III-3, III-4, III-5, IV-1, and a married-in II-2a) (Fig. 1). Two ancestral individuals were obligate variant carriers of unknown disease status: II-1 and II-2. Lymphoblastoid cell lines (LCLs) were also available from the same eight individuals established by ECACC (HPA, Porton). Both affected individuals had disease onset in their 60s, with a disease duration ranging from two to four years until death.

SALS samples: Spinal cord post-mortem tissue samples stored at −80°C were collected. Samples were taken from the lumbar regions (L3 to L5) of 49 individuals, of which there were 32 SALS cases and 17 controls. The median age at death was 68 years for patients with SALS and 66 years for controls. The median post-mortem delay in the sample collection was 14 hours for SALS cases and 11 hours for controls (Supplementary Table 4). The pathological characteristics of these tissue samples were described in detail in our previous work (Paul et al., 2014).

### Cell culture

We grew Epstein-Barr Virus (EBV)-transformed B-cell lines, lymphoblastoid cell lines (LCLs), in RPMI 1640 medium (Gibco) with 10% foetal bovine serum (Sigma-Aldrich), 1% glutamax (Gibco), 1% penicillin/streptomycin (Gibco) at 37°C, 5% CO_2_ until an appropriate number of cells was obtained. Cells were counted, and 1 ml of culture medium containing about 3 x 10^6^ cells was transferred into each well of a 24-well plate for subsequent drug treatment. We used the following drugs: tunicamycin (2 µg/ml) (Sigma-Aldrich), thapsigargin (0.5 µM) (Sigma-Aldrich), rapamycin (0.2 µM) (Sigma-Aldrich), and tertiary-butyl hydroperoxide (50 µM) (Sigma-Aldrich). The drugs were diluted with dimethyl sulfoxide (DMSO). After 24 hours of incubation at 37°C and 5% CO2, the cells were treated for 6 hours with thapsigargin, 24 hours with tunicamycin, and 24 hours with rapamycin. The experimental controls included wells containing either medium alone or medium supplemented with 0.4% DMSO, which matched the solvent conditions used for the drug treatments. The duration of DMSO exposure was equivalent to that of the corresponding drug treatment. Cold phosphate-buffered saline (PBS) was added to each well to inhibit the drug treatment. The cells were transferred into tubes and centrifuged for 5 min at 2000g. The supernatants were discarded, and the pellets were stored at −80°C for DNA, RNA, or protein extraction.

### DNA extraction and Sanger sequencing

Genomic DNA was extracted from whole blood or lymphoblastoid cell lines using the QIAamp® DNA Midi kit (Qiagen, UK) according to the manufacturer’s protocol. The extracted DNA was quantified using an ND-1000 spectrophotometer (NanoDrop®). The DNA had a 260/280 absorbance ratio of 1.8 or higher. For PCR amplification of *RUBCN*, we used forward primer AGGAGAATGCCCACTTCAGC and reverse primer CAGGTTCTTGGTGCGGATTT. For PCR amplification of *KANK1*, we used forward primer GGTTTGAGAAACCCAACATGGC and reverse primer GTGGAGGCGGAAAGGATGAA. Primers were designed using Primer3 software. The purity and yield of the PCR products were confirmed using agarose gel electrophoresis. The PCR products were treated with Exonuclease I and Shrimp Alkaline Phosphatase for the removal of unused primers and deoxynucleotide triphosphates (dNTPs). The PCR products were sequenced using Sanger sequencing at the MRC CSC Genomic Core Facility of Imperial College London. DNA sequence files were analysed using CodonCode aligner software.

### RNA extraction and RNA quality assessment

RNA was extracted from the frozen spinal cord post-mortem tissue samples and lymphoblastoid cell lines using the Direct-zol^TM^ RNA miniprep kit (Zymo Research, USA) according to the manufacturer’s protocol. Extracted RNA was eluted in 40 µl of DNase/RNase-free water. RNA was quantified using an ND-1000 spectrophotometer (NanoDrop®) and stored at −80°C. The 260/280 absorbance ratios ranged from 1.8 to 1.9 for all tissue samples. We measured the pH of the spinal cord tissues from SALS cases and controls. The median value of pH was 6.3 for SALS cases and 6.4 for controls (Anagnostou et al., 2010). The RNA integrity of the extracts was assessed using the Agilent 2100 Bioanalyzer system in combination with the RNA 6000 LabChip®. Total RNAs with 28S:18S rRNA ratios >1.0 were selected for further study (Montibeller & de Belleroche, 2018).

### cDNA synthesis and qPCR

cDNA was synthesised from RNA extracts using a cloned Avian Myeloblastosis Virus (AMV) cDNA synthesis kit (Invitrogen). cDNA was stored at −20°C for further use. Primers were designed using Primer3 software to amplify the target sequence (Supplementary Table 5).

qPCR reactions were prepared using PowerUp™ SYBR^®^ Green Master Mix (ThermoFisher Scientific) and 6 μM of primers. ROX dye was also included for signal normalisation to reduce variability between technical replicates. Amplification was performed using the Stratagene MX3000p qPCR system. The following cycling conditions were applied: initial denaturation at 95°C for 10 min, followed by 35 cycles of denaturation at 95 °C for 30 s, annealing at an optimized temperature for 30 s, and extension at 72 °C for 45 s. The final cycle included denaturation at 95 °C for 1 min, annealing at an optimized temperature for 30 s, and extension at 72 °C for 45 s. All the amplified qPCR products were analysed by observing thermal dissociation curves in addition to gel electrophoresis to confirm the purity and size of the products.

### FITC annexin V assay

For FITC annexin V assays, we grew the LCLs as described above. Cells were counted, and 1ml of culture medium containing 3 x 10^6^ cells was added to each well of a 24-well plate. The cells were seeded for approximately 24 hours, followed by drug treatment with tunicamycin, thapsigargin, and rapamycin, as mentioned earlier, along with the corresponding vehicle controls. The cells were also treated with staurosporine (5 uM) (Sigma-Aldrich) for 4 hours, along with the corresponding vehicle control. Staurosporine was used as a positive control for the apoptotic assay. Drug treatment was inhibited by adding cold phosphate buffer saline (PBS) to each well. The cells were subsequently stained for the assay using a FITC Annexin V Apoptosis Detection kit 1 (BD Biosciences) according to the manufacturer’s protocol. Cells were sorted using a two-laser FACSCalibur Flow Cytometer (BD Biosciences) to detect apoptosis. The data obtained was analysed using FlowJo 10.1 software.

### Variant Data and Statistical Analysis

An Imperial College cohort of 13 index cases underwent exome sequencing as part of an international consortium study (Smith et al., 2014), including a member of the British ALS kindred SM133, in which multiple individuals were affected by the disease. A missense variant was identified in both *RUBCN* and the *KANK1* genes in two cousins with ALS from the SM133 kindred (Fig. 1). The genotypes of the variants were subsequently determined by Sanger sequencing in eight family members or inferred from the three obligate carriers of this kindred.

In a combined control dataset comprising 53,424 individuals (106,848 chromosomes), *RUBCN* SNV were absent, and *KANK1* SNV were identified only once, indicating that these variants are ultra-rare. The combined control dataset consisted of 51,592 non-Finnish Europeans who had not been ascertained for a neurological condition (gnomad.broadinstitute.org, v3.1.2) 27, and 1,832 ALS controls from the MinE project, who were also of European ancestry (databrowser.projectmine.com) (van der Spek et al., 2019).

To assess the association of the two SNVs segregating in the family with ALS, the cosegregation of candidate variants with the disease was evaluated using the full-likelihood Bayes factor (FLB) method described by Thompson et al. (2003) (Thompson et al., 2003). This approach was chosen because it accounts for the pedigree structure, incomplete penetrance, phenocopies, and individuals with unknown phenotype or genotype, providing a more accurate assessment than meiosis counting methods (Jarvik & Browning, 2016). The FLB computes the ratio of the probability of the observed pedigree phenotype and genotype data under a model in which the variant is causal (H1) to the probability under a model in which the variant is not causal (H0), integrating over all unobserved genotypes using the Elston-Stewart peeling algorithm (Elston & Stewart, 2008). FLB calculations were implemented using the segregatr R package (github.com/magnusdv/segregatr) (Ratajska et al., 2023; Vigeland, 2021). Pedigree objects were constructed using the pedtools package (Vigeland, 2021) and all analyses were performed in R version 4.5.2. The proband was specified as the index affected carrier to implement ascertainment correction (Thompson et al., 2003). A phenocopy rate of 0.08 was applied (Turner & Talbot, 2013) as well as a missense variant allele frequency of < 0.001% in the 53,424 control sets noted above. FLB values were compared to the standards and guidelines of the American College of Medical Genetics and Genomics and the Association for Molecular Pathology (ACMG/AMP; Richards et al. 2015), for which single-family thresholds (Jarvik & Browning, 2016) are as follows: FLB ≥ 4.33 corresponds to PP1_Supporting; FLB ≥ 18.7 corresponds to PP1_Moderate; FLB ≥ 350 corresponds to PP1_Strong. Since all affected variant carriers were male and all unaffected variant carriers were female, a sex-stratified liability test was also implemented in segregatr to estimate the relationship between FLB and a grid of male and female penetrance values ranging from 0.01 to 0.99.

Rare variant collapsing analysis was applied to regional rare missense SNVs within the *RUBCN* and *KANK1* genes in ALS cases and controls. The former were retrieved from the MinE database of 4,366 ALS cases as well as 3,317 cases of ALSdb (github.com/igm-team/alsdb). In a review of sequence-based association studies of three disease groups, Povysil et al. consistently found that the greatest disease risk was associated with “ultra-rare” missense variants, defined by a minor allele frequency (MAF) < 0.001% and predicted deleteriousness; a similar association was also reported for quantitative traits (Povysil et al., 2019). Therefore, ultra-rare variants were selected with an MAF < 0.001% in 106,848 chromosomes from the gnomAD and MinE control datasets. All such variants were further filtered for pathogenicity, showing a CADD (Combined Annotation Dependent Depletion with logarithmic transformation) PHRED score ≥ 20 (cadd.gs.washington.edu/snv) (Rentzsch et al., 2019). Region-based tests of association for such variants with ALS in the above datasets were estimated using a hybrid burden and sequence kernel association test (optimized SKAT-O; CRAN.R-project.org/package=SKAT) for optimal power across different genetic architectures. For each alternate allele included in the aggregate variant set, the corresponding reference-allele entry in the input file was the number of successfully sequenced chromosomes carrying that specific reference allele, rather than the maximum number of chromosomes subjected to sequencing in the dataset. Consequently, the reference-allele count varied across variants according to the number of chromosomes with valid sequence data at that position. For example, for the control Ala74Val variant the alternate allele was observed once among 106,252 successfully evaluated chromosomes (corresponding to 53,126 individuals), out of a maximum dataset size of 53,205 individuals. The variant was therefore represented as an alternate-allele count of 1 with a variant-specific reference-allele count of 106,252. We used the ‘SKATBinary_Robust’ function to adjust for unbalanced case-control ratios (Zhao et al., 2020). For whole-exome sequencing (WES), Fadista, Manning et al. recommended an allele frequency-adjusted threshold of significance of P< 2.5×10^-7^ (Fadista et al., 2016).

Gene expression levels were normalised against house-keeping genes, *GAPDH* or *ACTB,* using transcript values of ΔCt =(Ct^gene^ ^of^ ^interest^ – Ct^housekeeping^ ^gene^). ΔCt, also called ΔCq, can be expressed as 2^-ΔCt^ = fold change (FC) values. Negative ΔCt values were plotted in graphs because an increase in transcript levels may be shown as an upward shift in the ordinates. Statistical analysis of gene expression was performed using GraphPad Prism software. The effect of drugs was analysed using the paired Student’s t-test and the effect of the mutation was analysed using the unpaired Student’s t-test (parametric) or Mann-Whitney U test (non-parametric). The effect of the mutation together with the drug treatment on cell survival was analysed using two-way ANOVA with a Bonferroni post-hoc test. Results were given as mean ± Standard error of the mean (SEM).

## Data Availability

All data produced in the present study are available upon reasonable request to the authors.

Summary statistics, including burden test results and cosegregation (FLB) analyses, are provided in the article and Supplementary Information. Individual-level genomic and pedigree data are not publicly available because they contain information that could compromise the privacy and identifiability of study participants and their family members, consistent with the terms of the informed consent and ethics approval under which the data were collected.

## Acknowledgements

This work is dedicated to the memory of Jacqueline de Belleroche, an intelligent scientist with passion and dedication who has contributed to scientific progress in the study of ALS. She was a mentor to all of us in her lab, and she will always be. We are grateful to the tissue donors and their relatives involved in this research through the Imperial College ALS Tissue Bank. We thank Luigi Montibeller and Maria Teresa Cencioni for their support of this study. We are grateful to Imperial College London for the award of President’s PhD scholarship to Midhat Salman, to Majlis Amanah Rakyat (MARA) Malaysia for the award of PhD scholarship to Intan Bakhtiar, to the MRC and MND associations for funding the project.

## Author contributions

M.S. performed the validation and functional characterisation of the p.C517F *RUBCN* and contributed to manuscript preparation. I.B. performed the validation and functional characterisation of the p.P1332A *KANK1* and contributed to manuscript preparation. A.M. performed genetic analysis and functional associations of both gene variants and contributed to manuscript preparation. A.R. performed a subset of the functional characterisation of p.C517F *RUBCN* and prepared figure 8. B.S., S.T., C.H. generated and analysed exome and genome data. B.S. and S.T. contributed to manuscript preparation. M.S., I.B., A.M., and J.D.B. designed this study.

