## Supplementary data for "Identification of co-segregating *RUBCN* and *KANK1* mutations in a UK amyotrophic lateral sclerosis kindred"

**Figures**

**A)**

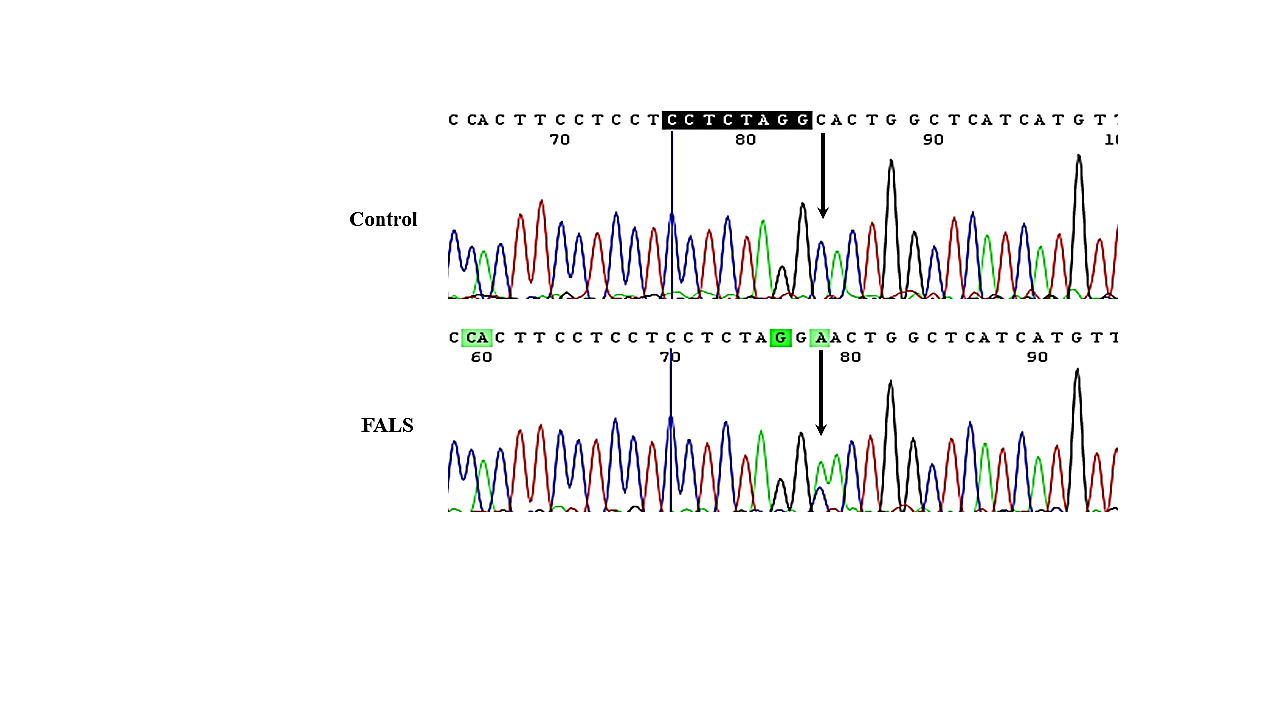

**B)**

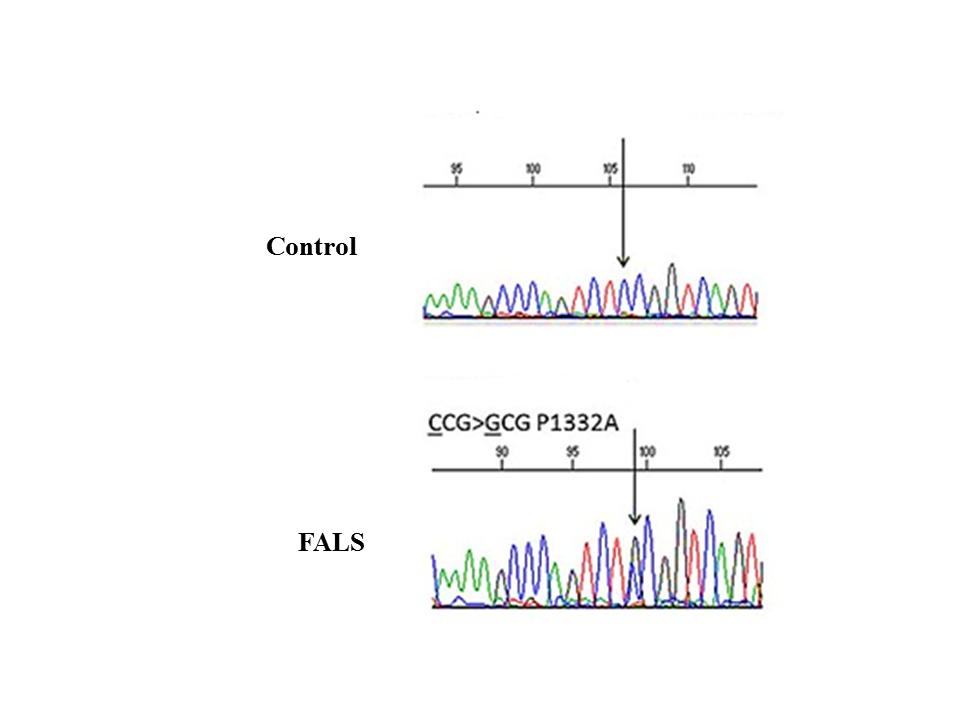

**Supplementary Figure 1. Chromatograms of *RUBCN* and *KANK1* sequences indicating the nucleotide transversion in FALS case compared to a control.** The chromatograms shown in **A)** indicate the difference between the DNA sequence of *RUBCN* in a control and a FALS case. The position of a heterozygous transversion, C to A, in the reverse strand of FALS case for *RUBCN* c.1550G>T (C517F) compared to a control has been indicated by arrows. The chromatograms shown in **B)** indicate the difference between the DNA sequence of *KANK1* in a control and a FALS case. The position of a heterozygous transversion, C to G, in *KANK1* c.3994C>G (P1332A) compared to a control has been indicated by arrows.

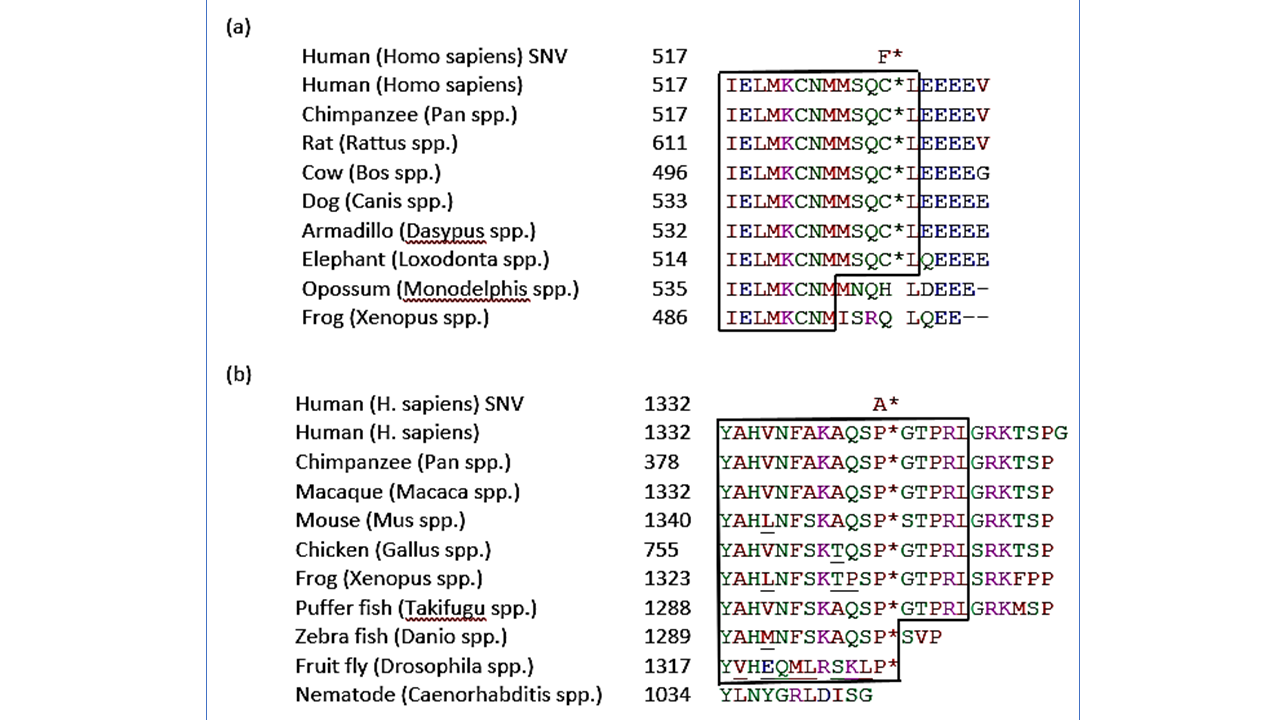

Supplementary Figure 2. Flanking sequence of missense variants in Rubicon and Kank1. (a) p.C517F (wild-type F* and variant C*) in human Rubicon and its orthologs as well as (b) p.P1332A (wild-type A* and variant P*) in human Kank1 and its orthologs. Boxes show conserved regions of identical amino acids except for underlined amino acids that are not identical to other residues in the same column but show a homologous substitution when the colour is shared.

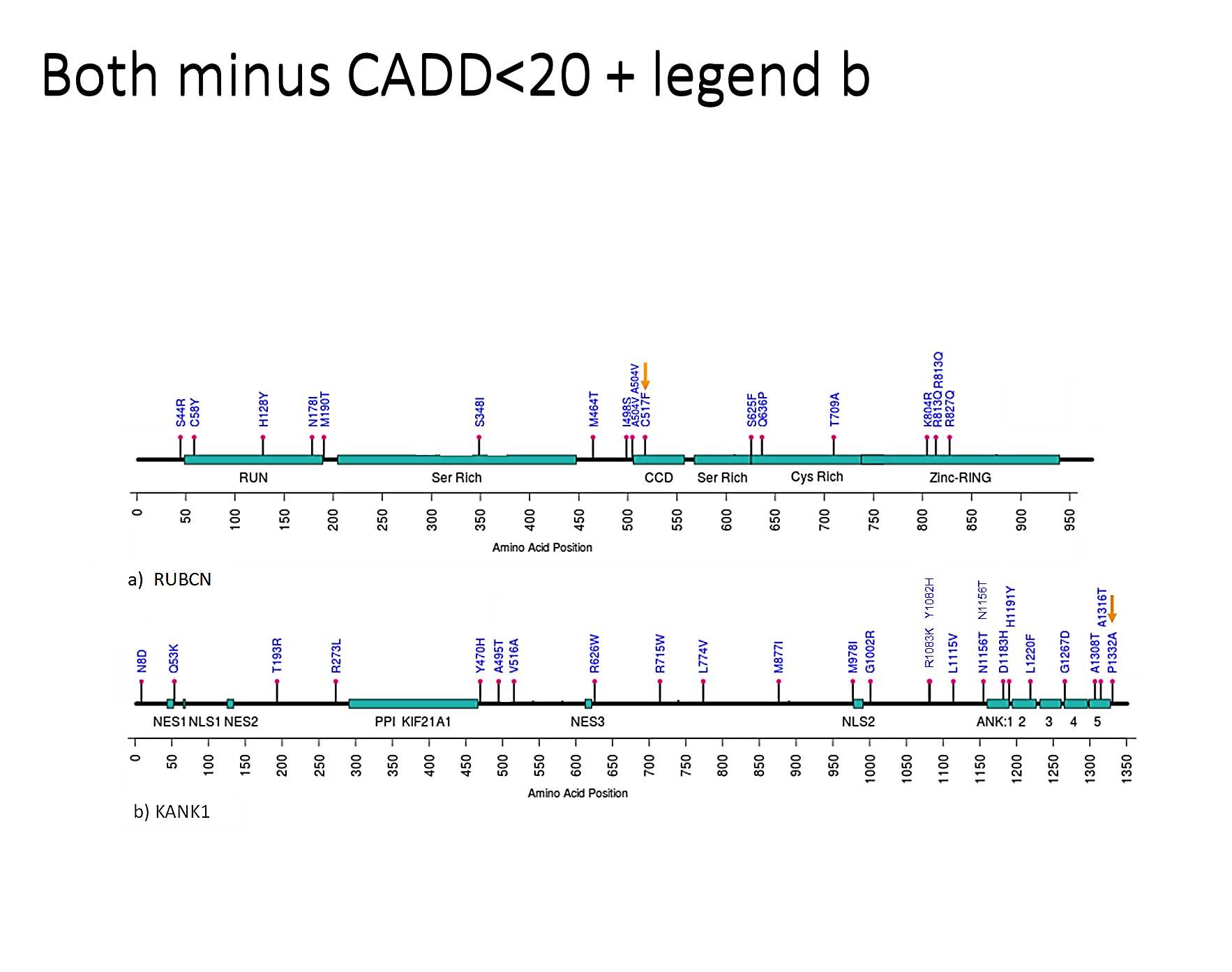

Supplementary Figure 3. Missense SNVs of *RUBCN* and *KANK1*.

Missense SNVs found in 7696 ALS patients from the MinE and ALSDB3 databases were selected for an ultra-rare frequency <0.001% in 53,424 neurological controls (MinE controls and the gnomAD non-Finnish European individuals that were diagnosed not to have a neurological disease). **(a)** Ultra-rare ALS SNVs in *RUBCN*-202 showing C517F (yellow arrow) and a further 16 variants with a CADD score >20, including the two doubletons at A504V, R813Q. **(b)** 25 ultra-rare ALS SNVs in *KANK1*-206 with a CADD score >20, showing P1332A (yellow arrow) and the doubleton at N1156T. The 5 and 20 rare control variants found respectively in *RUBCN* and *KANK1* were selected using the same criteria but are not depicted. Abbreviations: Ankyrin domain (ANK) repeats; Coiled-Coil Domains (CCD); Nuclear Export Signal 1 (NES): Nuclear Localization Signal 1 (NLS); Protein-Protein Interaction (PPI); RPIP8, Unc-14 And NESCA domains (RUN). The orange arrows show the two SNVs found in kindred SM133.

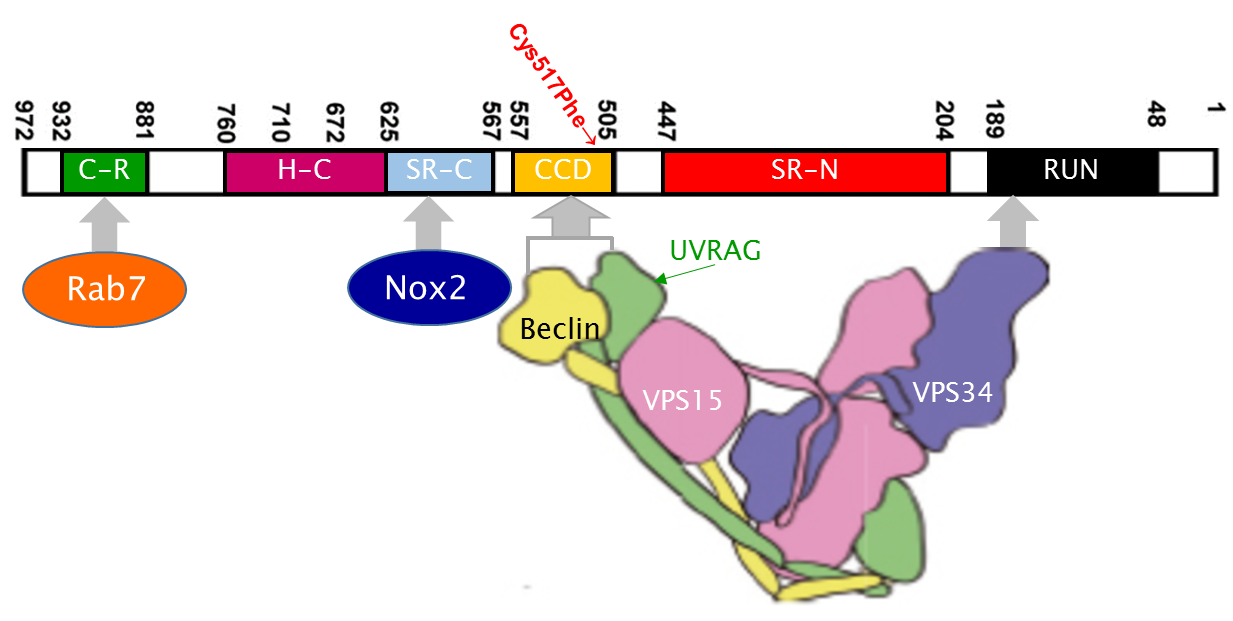

**N-terminus**

**C-terminus**

Supplementary Figure 4. Rubicon interacts with proteins from several pathways.

The C-terminal half interacts with both Rab7 and Nox2, where the p.C517F mutation (red arrow) was associated with a decreased expression of both Rab7 and Nox2 transcript levels. Along the N-terminal domains, Rubicon interacts with components of the PI3KC3 complex: Vps34, UVRAG and Beclin-1. Their expression levels showed no association with this mutation. There are multiple interactions present between the PI3KC3 components as shown by the intertwined subunits (Rostislavleva et al., 2015). The numbering from right to left refers to the amino acid position from N- to the C-terminus. RUN: Run domain; SR-N: serine rich-N terminus domain; CCD: coiled-coil domain; SR-C: serine rich-C terminus domain; H-C: helix-coil-rich domain; C-R: cysteine-rich domain.

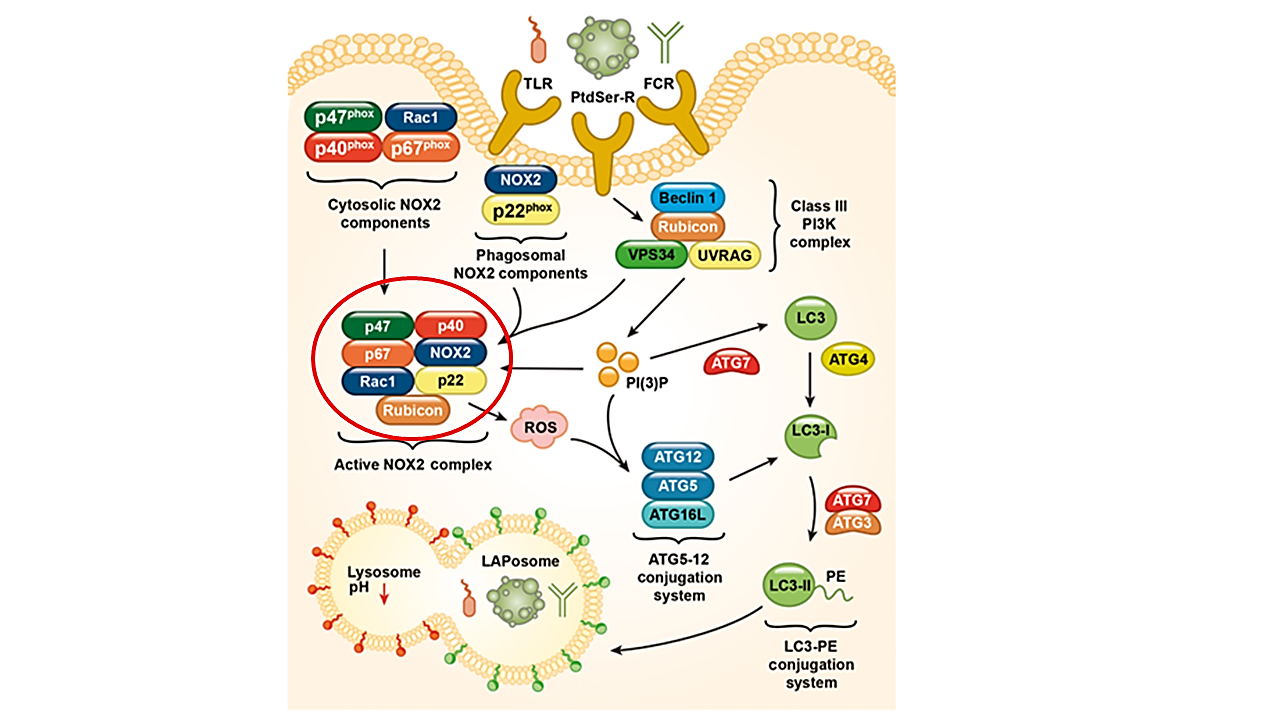

**Supplementary figure 5. LC3-associated phagocytosis pathway.**

When the stimuli that activate cell surface receptors (Toll-like receptors, TLR; Fc receptors, FCR; and phosphatidylserine receptors, PtdSer-R) are taken up by the cell, components of the LC3-associated phagocytosis (LAP) pathway are recruited to the LAPosome. This results in the interaction of the Rubicon containing PI3KC3 complex with the LAPosome and production of PI(3)P. In turn, PI(3)P performs two roles: it both activates and stabilises the NOX2 complex (cytosolic NOX2 components join phagosomal NOX2 components) for ROS generation and recruits the downstream ATG5-12 and LC3-PE conjugation systems. Additionally necessary for the stability of NOX2 complex is Rubicon. The subsequent anchoring of LC3-II on the LAPosome membrane by both PI(3)P and ROS causes the maturation of LAPosomes to fuse with lysosomes for the degradation of the cargo contained by the LAPosomes. Red circle indicates NOX2 protein whose mRNA expression was downregulated by the Cys517Phe *RUBCN* mutation. The decreased levels of NOX2 may decrease the level of reactive oxygen species (ROS) production and quash the noncanonical autophagy pathways such as LAP. Figure adapted from (Wong et al., 2018)

**Tables**

Supplementary Table 1. Pathogenicity prediction

| **Gene** | **Mutation** | **Pathogenicity Prediction** | | |
| --- | --- | --- | --- | --- |
|  |  | **PolyPhen 2 (max 1)** | **Mutation Taster (max 1)** | **CADD** |
| ***KANK1*** | p.Pro1332Ala | 0.997 | 0.999 | 25 |
|  |  | probably damaging | high probability of disease | top 0.32% of deleterious variants |
| ***RUBCN*** | p.Cys517Phe | 0.733 | 0.939 | 17.3 |
|  |  | possibly damaging | high probability of disease | top 1.9% of deleterious variants |

Pathogenicity prediction for p.Pro1332Ala and p.Cys517Phe with PolyPhen 2, Mutation Taster and Combined Annotation-Dependent Depletion (CADD).

Supplementary Table 2. Investigation of the effect of the mutations on cellular pathways under basal conditions at mRNA level.

| **Genes** | **Medium** | **DMSO**  **4 hr** | **DMSO**  **6 hr** | **DMSO**  **24 hr** |
| --- | --- | --- | --- | --- |
| ***RUBCN*** | ns | ns | ns | ns |
| ***KANK1*** | 0.0203↑ | - | - | - |
| ***LAMP2*** | ns | ns | ns | ns |
| ***VPS34*** | ns | ns | - | ns |
| ***UVRAG*** | ns | ns | - | ns |
| ***RAB7A*** | 0.0154↓ | ns | ns | ns |
| ***DNAJC10*** | ns | 0.0308↓ | - | ns |
| ***XBP1s*** | 0.006↑ | - | - | - |
| **NOX2** | ns | 0.0146↓ | - | ns |
| ***DNAJB9*** | ns | - | ns | ns |
| ***HSPA5*** | ns | - | ns | ns |
| ***SQSTM1*** | ns | - | ns | ns |

P-values obtained from unpaired t-tests investigating the effect of *RUBCN* and *KANK1* mutation on the autophagy/endosomal and LAP pathways in LCLs. LCLs were treated with 4 hours, 6 hours or 24 hours with the addition of vehicle, dimethyl sulfoxide (DMSO), depending on the corresponding drug treatment in the relevant experimental controls. The values of gene expression were normalised to the house-keeping gene β-actin. P-values indicate significant values with *p*<0.05 while ns indicates non-significant p-values. Arrows indicate an upregulation (↑) or downregulation (↓) in the expression of the gene.

**Supplementary Table 3. Investigation of the effect of drug treatments on cellular pathways at mRNA levels**

| **Pathway** | **Gene Transcript** | **Effect of Treatments on Transcript Levels in LCLs** | | | | | |
| --- | --- | --- | --- | --- | --- | --- | --- |
|  |  | **Rapamycin** | | **Tunicamycin** | | **Thapsigargin** | |
|  |  | **Wild type** | **Mutant** | **Wild type** | **Mutant** | **Wild type** | **Mutant** |
| Autophagy or LAP | *RUBCN* | 0.0073 | 0.0072 | Ns | ns | ns | Ns |
|  | *VPS34* | 0.0424 | ns | 0.0284 | ns | - | - |
|  | *UVRAG* | 0.0355 | - | Ns | ns | - | - |
|  | *NOX2* | ns | - | Ns | 0.0197 | - | - |
|  | *RAB7A* | 0.0144 | ns | Ns | ns | ns | Ns |
| Lysosome Trafficking | *LAMP2* | ns | ns | 0.017 | 0.038 | ns | Ns |
| ER Stress Response | *HSPA5* | ns | ns | 0.016 | 0.002 | ns | 0. 021 |
|  | *DNAJC10 / PDIA19* | 0.0168 | ns | Ns | ns | - | - |
|  | *DNAJB9* | ns | ns | Ns | ns | ns | 0.0007 |
|  | *XBP1* | ns | ns | 0.0264 | 0.0289 | - | Ns |

The effects of drug treatment on the transcript levels in wild type and mutant lymphoblastoid cell lines (LCLs) (*RUBCN* C517F and *KANK1* P1332A in members of kindred SM133) involved in the pathways of LAP, lysosome trafficking and ER-stress response. Differences between the mutant and wild type due to rapamycin treatment showed significant downregulation of *VPS34, RAB7A* and *DNAJC10* in the wild type, compared to no change in the corresponding mutant cell lines. Abbreviations: ‘ns’ not significant; ‘-’ not determined; ‘🡹’transcript levels significantly up- or ‘🡻’ downregulated (*p* < 0.05).

**Supplementary Table 4. Spinal cord tissue samples from SALS cases and controls.**

| **Group** | ***n*** | **Age at death (years)**  **Median (range)** | **Post-mortem delay (hours)**  **Median range** |
| --- | --- | --- | --- |
| SALS | 32 (21 males, 11 females) | 68 (24-85) | 14 (5-25) |
| Controls | 17 (12 males, 5 females) | 66 (20-91) | 11 (3-19) |

The controls and sporadic amyotrophic lateral sclerosis (SALS) cases included in the study along with their age at death and the post-mortem delay duration.

**Supplementary Table 5- Primers used for qPCR**

| **Gene** | **Primers** | **Product size (bp)** |
| --- | --- | --- |
| **Candidate genes** | | |
| *RUBCN* Both 972aa and 927aa transcripts | Forward: GAGAATGCCCACTTCAGCAT  Reverse: CAGGTTCTTGGTGCGGATTT | 180 |
| *RUBCN* 972aa transcript | Forward: GAATGGAGCTCGGAGGCGGCGA  Reverse: CGGATAAGCCCGTGATAGAG | 193 |
| *RUBCN* 927aa transcript | Forward: GCCCCAGGAATATCACCATT  Reverse: AGAGCTGGGTGTGCTGACTT | 122 |
| *KANK1* | Forward: GCACCCTGTCGTCTATCAACTC  Reverse: CTGCTGATTGGCTTTCCTTCT | 214 |
| *KIF21A* | Forward: GCAGAAGGGCAAGAGATTGGA  Reverse: TGCTTTCTCATATGCCTTTCTCCT | 183 |
| **Autophagy genes** | | |
| *RAB7A* | Forward: CACTCATGAACCAGTATGTG  Reverse: CATATCTGCATTGTGACTAGC | 118 |
| *LAMP2* | Forward: CGTTCTGGTCTGCCTAGTCC  Reverse: CAGTGCCATGGTCTGAAATG | 173 |
| *SQSTM1* | Forward: CAGTCCCTACAGATGCCAGA  Reverse: TCTGGGAGAGGGACTCAATC | 142 |
| *BECN1* | Forward: TGTCACCATCCAGGAACTCA  Reverse: CTGTTGGCACTTTCTGTGGA | 180 |
| *ATG14L* | Forward: ATGAGCGTCTGGCAAATCTT  Reverse: CCCATCGTCCTGAGAGGTAA | 192 |
| *UVRAG* | Forward: AGGGTTATTCAAATGCTCAG  Reverse: GTAGCAAAGAGAAGACATCG | 87 |
| *VPS34* | Forward: CTGTGCTGGATATTGCGTGATC  Reverse: AAGTCTATGTGGAAGAGTTTGCCT | 102 |
| **ER stress genes** | | |
| *XBP1 total* | Forward: GCTCAGACTGCCAGAGATCG  Reverse: TCTTCAGCAACCAGGGCATC | 188 |
| *XBP1 spliced* | Forward: AGAGTCTGATATCCTGTTGG  Reverse: AGTTCATTAATGGCTTCCAG | 189 |
| *HSPA5* | Forward: TCAAGTTCTTGCCGTTCAAGG  Reverse: AAATAAGCCTCAGCGGTTTCTT | 148 |
| *DNAJB9* | Forward: TTTCCAGACACGCCAGGATG  Reverse: GTCCTGCAGTGCTTGCTAGA | 195 |
| *DNAJC10* | Forward: CAGTGAAATATCATGGAGACAG  Reverse: ATTTCCTGTCCAAAGTTCTG | 95 |
| **Oxidative stress/ LC3-associated phagocytosis gene** | | |
| *NOX2* | Forward: AAGATCTACTTCTACTGGCTG  Reverse: AGATGTTGTAGCTGAGGAAG | 124 |
| **House-keeping genes** | | |
| *ACTB* | Forward: GACAACGGCTCCGGCATGTG  Reverse: CCTTCTGACCCATGCCCAC | 121 |
| *GAPDH* | Ready-made primer pair ordered from Primer design | 118 |
